# A Bayesian hierarchical approach to account for reporting uncertainty, variants of concern and vaccination coverage when estimating the effects of non-pharmaceutical interventions on the spread of infectious diseases

**DOI:** 10.1101/2022.06.20.22276652

**Authors:** Raphael Rehms, Nicole Ellenbach, Eva Rehfuess, Jacob Burns, Ulrich Mansmann, Sabine Hoffmann

## Abstract

Coronavirus disease (COVID-19) has highlighted both the shortcomings and value of modelling infectious diseases. Infectious disease models can serve as critical tools to predict the development of cases and associated healthcare demand and to determine the set of non-pharmaceutical interventions (NPI) that is most effective in slowing the spread of the infectious agent. Current approaches to estimate NPI effects typically focus on relatively short time periods and either on the number of reported cases, deaths, intensive care occupancy or hospital occupancy as a single indicator of disease transmission. In this work, we propose a Bayesian hierarchical model that integrates multiple outcomes and complementary sources of information in the estimation of the true and unknown number of infections while accounting for time-varying under-reporting and weekday-specific delays in reported cases and deaths, allowing us to estimate the number of infections on a daily basis rather than having to smooth the data. Using information from the entire course of the pandemic, we account for the spread of variants of concern, seasonality and vaccination coverage in the model. We implement a Markov Chain Monte Carlo algorithm to conduct Bayesian inference and estimate the effect of NPIs for 20 European countries. The approach shows good performance on simulated data and produces posterior predictions that show a good fit to reported cases, deaths, hospital and intensive care occupancy.

## 1 Introduction

The experience with coronavirus disease 2019 (COVID-19) during the past two years has underlined both the importance of and the challenges in the modelling of infectious disease. Infectious disease models can serve as critical tools to predict health care demand and to determine when and which non-pharmaceutical interventions (NPI) should be implemented to slow the spread of the infectious agent. However, the modeling of infectious disease is complicated by the fact that the main quantity of interest, i.e., the number of daily infections, is a latent variable that cannot be observed and therefore has to be estimated by using information on observable quantities. The number of reported cases, deaths and hospital occupancy all provide complementary, yet sometimes contradictory, information on the number of infections in a given geographical region. The number of reported cases is prone to under-reporting, as it depends both on testing capacity and the employed testing strategy (May, 2020). When changes in the testing strategy concur with the introduction or the relaxation of NPIs, they can create severe distortions in the estimation of NPI effects. Moreover, there is a delay in the reporting of new cases, which typically shows strong variations depending on the weekday. While modeling disease mortality, which is less prone to under-reporting, can avoid biases due to changes in the testing strategy, the time lag between infection and death may be highly variable, leading to a reduction of statistical power (Sharma et al., 2021). Focusing solely on disease mortality makes it difficult to predict health care demand in the future. A solution for this situation, in which we have several imperfect proxy variables for the number of infections, is to combine information on disease incidence, hospital occupancy and mortality in a common framework while explicitly accounting for time-varying under-reporting and reporting delays.

Bayesian hierarchical approaches can address this challenge by providing a coherent and flexible framework to integrate all available sources of information while accounting for different sources of uncertainty. By combining different submodels through conditional independence assumptions, it is possible to integrate mechanistic assumptions on disease dynamics and sub-models describing the relationship between the true (and unknown) number of infections and reported cases, deaths and hospital occupancy. Additionally, we can borrow information from other geographical regions to stabilize parameter estimates and to improve forecasts on future hospital demand. Current approaches to assess the effect of NPIs typically either focus on the number of deaths (Flaxman et al., 2020) or the number of cases (Dehning et al., 2020; Banholzer et al., 2020; Li et al., 2021; Islam et al., 2020). Unwin et al. (2020), Brauner et al. (2021) and Sharma et al. (2021) extend the semi-mechanistic Bayesian hierarchical model proposed by Flaxman et al. (2020) by including information on reported cases and deaths when inferring the number of new infections. However, these approaches typically only estimate NPI effects for short time periods because they do not explicitly account for differences in host susceptibility over time (due to vaccination or previous infection), seasonality, the prevalence of different variants of concern or time-varying under-reporting.

The aim of this work was to extend current Bayesian hierarchical approaches by integrating the available information on the number of reported cases, the number of deaths and hospital and intensive care unit (ICU) occupancy while accounting for under-reporting and reporting delays in the number of reported cases. We account for the influence of seasons, vaccination coverage and the prevalence of variants of concern as these factors can have a critical influence on the number of new infections and the relationship between infections and reported deaths and hospital occupancy. By doing so, it is possible to use data on almost the entire course of the pandemic in several countries rather than focusing on short time periods in a single country during which the variant, vaccination coverage and the testing strategy remained roughly constant. By allowing for weekday-specific delays in reported cases and deaths (that mainly arise due to reduced reporting during the weekend), we are not required to smooth the analyzed time series and can estimate the number of infections on a daily basis. We apply our approach to data on reported cases, deaths and hospital occupancy for COVID-19 from 20 European countries and investigate its performance both on simulated data and by assessing posterior predictions.

## 2 The model

In this section, we describe the Bayesian hierarchical model to estimate the effects of NPIs. The full model is depicted in Figure 1 as a Directed Acyclic Graph (DAG). At its core, the model treats the number of true and unknown infections at every time point and geographical region as a discrete latent variable. These infections are modeled using a *renewal model* which represents the contagion process of the disease. It describes the number of new infections at every time *t* ∈ {*t*_1_, *t*_2_, …, *T*} as a function of past infections while accounting for the generation time distribution, i.e., the time lag between a primary and secondary infection, and for a time-dependent instantaneous reproduction number. *t* represents an arbitrary unit of time, for example days. The renewal model can be seen as a more flexible version of the disease dynamics described in classical compartmental models for infectious disease (Wallinga and Lipsitch, 2007). We explain our model in two parts. First, we describe how we estimate the effects of NPIs given the true and unknown number of infections represented through the discrete latent variable while accounting for seasonality, vaccination coverage and variants of concern. Second, we describe how we infer the number of true and unknown infections (i.e., the values of the latent variable) based on the available information on reported cases, deaths and hospital and ICU occupancy.

**Figure 1:**
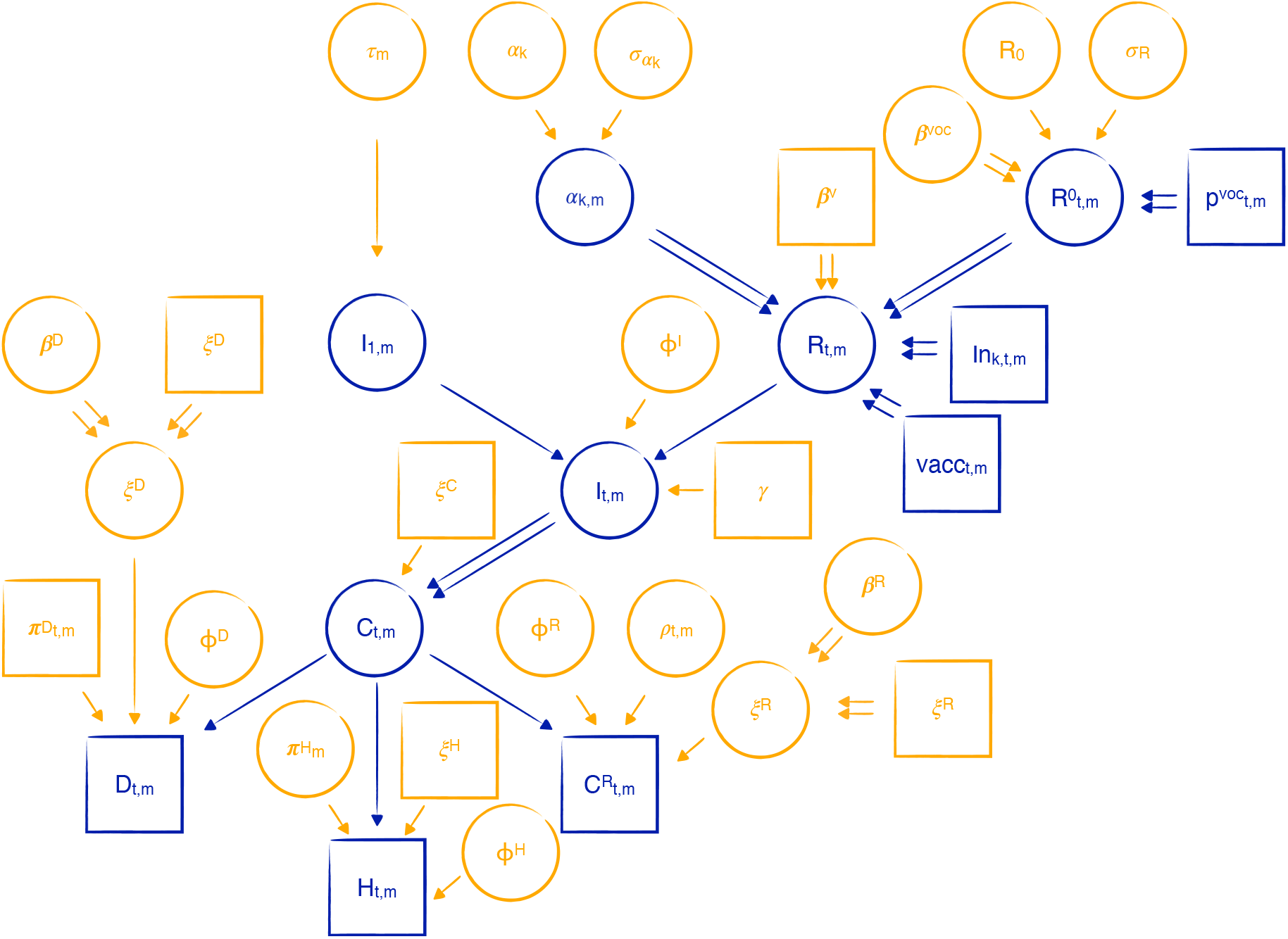
Directed Acyclic Graph (DAG) of the Bayesian hierarchical model. Quantities that are observed or assumed to be known are shown in squares and unknown quantities are shown in circles, single arrows indicate probabilistic dependencies and double arrows indicate deterministic dependencies. Parameters are highlighted in orange and variables are shown in blue. The model estimates the number of new infections *I*_*t,m*_ ∈ ℕ at every time *t* in geographical region *m* by using information on the number of deaths *D*_***t***\*,*m*_ ∈ ℕ, hospital occupancy *H*_***t***\*,*m*_ ∈ ℕ and reported cases 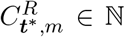 observed on the collection of future time points ***t***^*^ in this geographical region. The true and unknown number of cases *C*_*t,m*_ ∈ ℕ on day *t* in geographical region *m* is linked to the number of infections *I*_*t,m*_ through a deterministic function (indicated through a double arrow) that shifts the number of infections by the incubation period distribution *ξ*^*C*^. The two distributions *ξ*^*R*^ and *ξ*^*D*^ describe the time between the onset of symptoms and reporting as a case and death, respectively. *ξ*^*H*^ is the conditional probability of being in hospital *x* days after symptom onset. The parameters 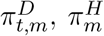 and *ρ*_*t,m*_ denote the probability of dying (i.e., the infection fatality rate), the probability of being hospitalized and the probability of being reported (i.e., the case detection ratio), respectively. For ICU occupancies, all parameters are defined in the same manner as for hospital occupancy, so we omit information on ICU in the DAG for sake of clarity.

### 2.1 Estimation of NPIs

#### Renewal Equation

To model the dynamics of the infectious disease, we use a renewal equation which reflects the spreading of the infectious agent among susceptibles:

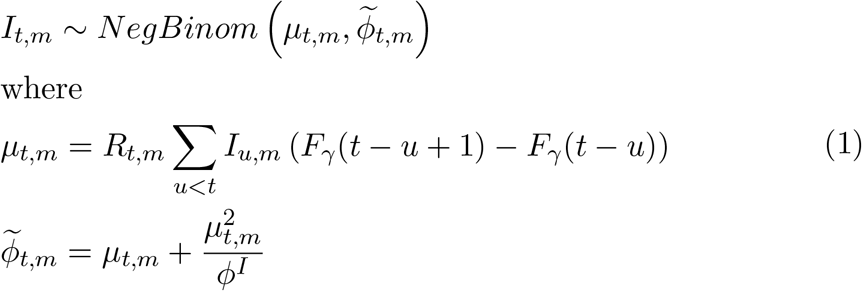

Equation (1) describes the number of infected individuals *I*_*t,m*_ at each time point *t* in geographical region *m* ∈ {*m*_1_, …, *M*} as a function of past infections, the instantaneous reproduction number *R*_*t,m*_ (see next subsection for more details) and the generation time distribution. In equation (1), the number of infections *I*_*t,m*_ is the sum of the previous infections on the *t* − 1 days before *t* weighted by the corresponding probability mass of the discretized generation time distribution *F*_*γ*_(*t*−*u*+1)−*F*_*γ*_(*t*−*u*) multiplied by the local instantaneous reproduction number *R*_*t,m*_ at time *t*. Applying the renewal equation to past infections yields the current number of infections *I*_*t,m*_ (see for instance Fraser et al. (2009)). To reflect uncertainty in the renewal model, we consider infections at time *t* in geographical region *m* to follow a Negative Binomial distribution with expectation *μ*_*t,m*_ and an overdispersion parameter *ϕ*^*I*^. For *ϕ*^*I*^ → ∞, *I*_*t,m*_ would follow a Poisson distribution with no overdispersion.

We seed the model for the first day *I*_1,*m*_ in each geographical region *m* through a Negative Binomial distribution with mean parameter *τ*_*m*_:

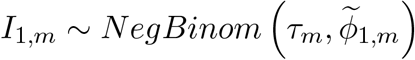

where we assume a hierarchical model for *τ*_*m*_, i.e., each *τ*_*m*_ follows a truncated normal distribution around a common parameter *τ* which follows a Gamma distribution with shape *a*_*τ*_ and scale *b*_*τ*_.

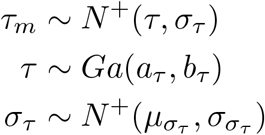

#### Effects on the reproduction number

Besides past infections and the generation time distribution, the renewal equation also includes the instantaneous reproduction number *R*_*t,m*_. We define *R*_*t,m*_ as the product of three factors: the basic reproduction number 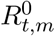 (which may also be time dependent due to the influence of different variants of concern), the effect of NPIs and two correction factors 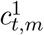 and 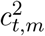:

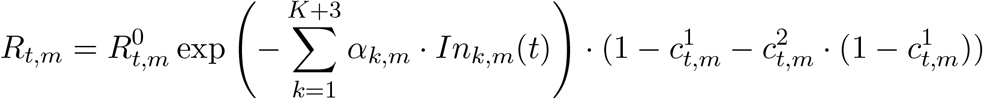

with

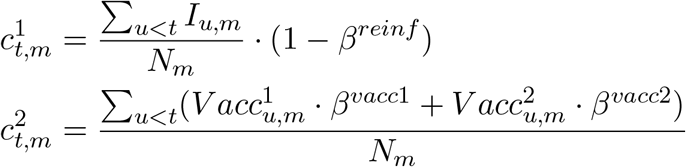

The correction factors 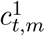and 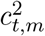reduce the number of susceptibles in the population: 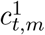corrects for already infected and possibly recovered individuals. Since an infection may not guarantee protection against the infectious agent, we include a parameter *β*^*reinf*^ giving the probability of reinfection. The term 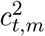 corrects for vaccination coverage at time *t* in geographical region *m*. Here, *β*^*vacc*1^ and *β*^*vacc*2^ represent the probability of infection after a first and second vaccination. 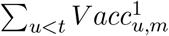 and 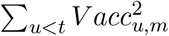 are the number of vaccinated individuals in the population at time *t* and geographical region *m*.

To estimate the effect of NPIs, we include a further factor in the instantaneous reproduction number. Here, the effect of *K* NPIs *α*_*k,m*_ is modeled inside an exponential function with an corresponding covariate *In*_*k,m*_(*t*) which is an indicator taking a value of 1 if the *k*-th NPI is active at time *t* in geographical region *m* and 0 otherwise. Besides NPIs, we also include three further factors to account for the effect of seasons (choosing summer as reference category, resulting in *K*+3 indicator variables). Since it is reasonable to assume variations in the effectiveness of NPIs and seasons between different geographical regions, we allow for an individual effect using a hierarchical structure. This hierarchical structure makes it possible to share information between regions to infer an overall effect of the NPIs while allowing to estimate individual effects that are specific to geographic regions to uncover variation in these effects.

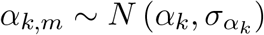

The basic reproduction number may vary over time due to the occurrence of new variants that modify 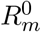. We propose a convex combination to construct a time-dependent basic reproduction number 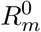. For the application to COVID-19, we account for two variants of concern yielding the following formula:

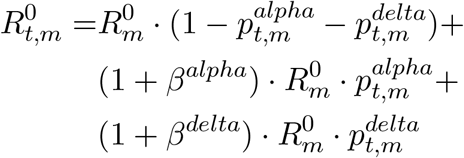

Here 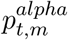 and 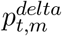 are the prevalence of the alpha (B.1.1.7) and delta (B.1.617.2) variants, respectively, at each time *t* in geographical region *m*. The two unknown parameters *β*^*alpha*^ and *β*^*delta*^ represent the over-contagiousness of these variants compared to the wild type. We obtain a time variant reproduction number by taking the reproduction number of the original wild type as basis and multiplying it with (1+*β*^*alpha*^) and (1+*β*^*delta*^) which accounts for the effect of these subsequent variants.

Since each geographical region may have its own characteristics, we allow for variation in the basic reproduction number. We therefore assume reproduction numbers 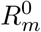 that are specific to geographical region *m* that are again modeled in a hierarchical manner with common mean *R*_0_:

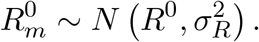

### 2.2 Inferring the number of infections

To estimate the number of infections on day *t* in geographical region *m*, we use information on four observed time series where each of them is linked through a submodel to this latent variable: the reporting model, the death model and two hospitalization models (normal beds and ICU).

#### Disease model

Before linking the number of infections to the observable time series, we define a second latent variable, the number of cases *C*_*t,m*_ in geographical region *m* with symptom onset on day *t*, which is simply a deterministic function of the number of infections *I*_*t,m*_ occurring until time *t*:

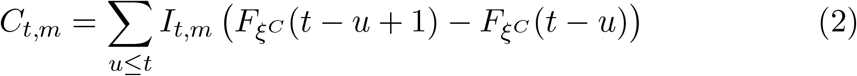

where 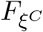 is the cumulative distribution function of the incubation period. Through this relation, the number of cases becomes a deterministic function of the number of infections which is shifted by the incubation time distribution. As the disease model described in equation (2) produces values for *C*_*t,m*_ that are possibly not integers, we apply a rounding scheme which both ensures integer values for the cases and that the total number of cases equals the total number of infections. If *C*_*t,m*_ is not an integer, we round it to a full number while keeping the remaining decimals to add or subtract them to *C*_*t*+1,*m*_, i.e., the number of cases on the next day in geographical region *m* ^1^.

### 2.3 Reporting model

In the reporting model, we describe the association between the number of cases *C*_*t,m*_ with symptom onset on day *t* and reported cases. By accounting for time-varying under-reporting and weekday specific reporting delay, the reporting model can give insight into the discrepancies between the true dynamics of the disease and the official information recorded by health authorities.

#### Time-varying under-reporting

Following work by Fraser et al. (2009) and Azmon et al. (2014), we account for discrepancies in the number of reported and actual cases. Since the testing behavior varies over time, we propose a time-dependent case detection ratio *ρ*_*t,m*_ in each geographical region *m*. We assume the reporting rate to be a piece-wise constant function with predefined cut-points. These cut-points can be predefined as time-points at which structural changes in testing strategies or procedures occurred.

#### Reporting delay

Many local authorities follow a protocol or rules in their daily data gathering process, which lead to strong weekly variations in the reported data (Günther et al., 2021). We extend the time-varying under-reporting by including a weekday specific reporting delay. We do so by using a discrete convolution in the same manner as for the generation time distribution in the renewal model while accounting for time-varying under-reporting:

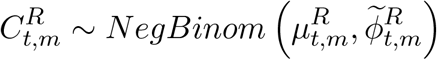

where

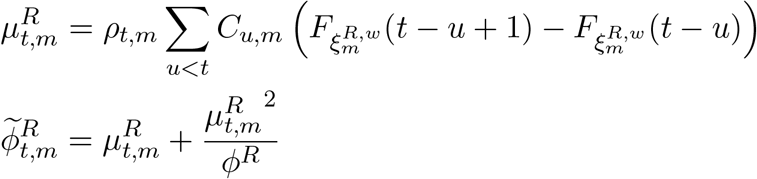

Here, 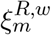 is the reporting delay distribution for a specific weekday *w* in geographical region *m*. The expected number of reported cases on day *t* is a sum of all true cases occurring on some day *u* before day *t* weighted by their probability of being reported after *t* − *u* days and multiplied by the time-specific under-reporting rate *ρ*_*t,m*_. Here *ρ*_*t,m*_ is described through apiece-wise constant function.

The reporting model requires detailed knowledge about the distribution of the delay between symptom onset and day of reporting by the health authorities in a geographical region *m*. Since it is very difficult to obtain this information for each region, we suggest using a general reporting delay distribution and introducing parameters 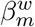 to adapt this distribution for specific weekdays *w* for each geographical region *m*. These parameters inflate or deflate the probability mass on a specific day of the week to reflect the systematic discrepancies in the observed data 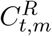 and the expected number of reported cases. 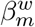 affects the distribution as follows: Given the variable 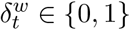 indicating that day *t* is weekday *w* in a (weekday specific) baseline distribution 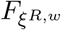, we obtain a modified version of 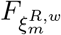 by calculating 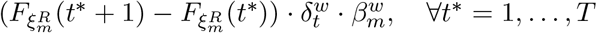 and re-normalizing it afterwards. As a consequence, we can account for weekly seasonality effects that are specific to each geographical region.

### 2.4 Death model

Following Flaxman et al. (2020), we describe the number of deaths *D*_*t,m*_ occurring on day *t* in geographical region *m* as a function of the number of true cases with disease onset prior to *t*. In this death model, *D*_*t,m*_ is described by a Negative Binomial distribution with expected value equal to the sum of the number of true cases with disease onset at time *t* − *u*, weighted by the probability of dying on the *u*-th day after the onset of symptoms. This latter probability can be obtained by discretizing the probability distribution describing the time until death for patients who died, i.e., 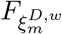, and multiplying by the infection fatality rate (IFR) 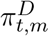, i.e., the probability of dying for an infected individual where this rate can depend on day *t* and geographical region *m*. Similarly to 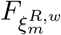 in the reporting model, 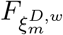 accounts for weekday effects which can be specific to geographical region *m* to account for differences in reporting.

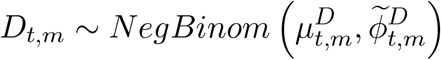

where

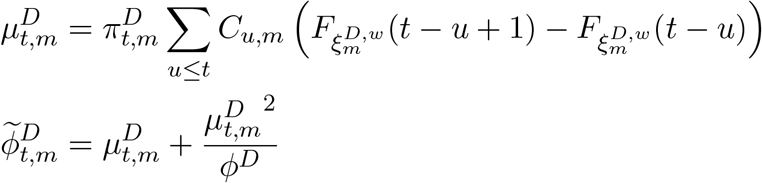

We use a region-specific IFR 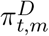, since the age structure of the geographical regions may be very different leading to variations in the severity of the disease. Since the model aims to use a long observation window, we have to reflect additional effects on the IFR, in particular concerning the effect of vaccinations and the effect of new variants of the disease which can become more or less prevalent over time. The IFR is therefore a time- and location-dependent quantity. Details on how we adapt the IFR in the context of our application to COVID-19 (section 5) can be found in sectionC.3 in the supplementary material.

### 2.5 Hospitalization models

Since many countries provide information on hospital (normal beds) and ICU occupancy, we integrate these two additional sources of information through two hospitalization models whose structure is similar to that of the death model:

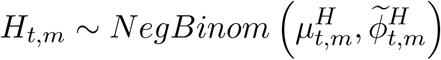

where

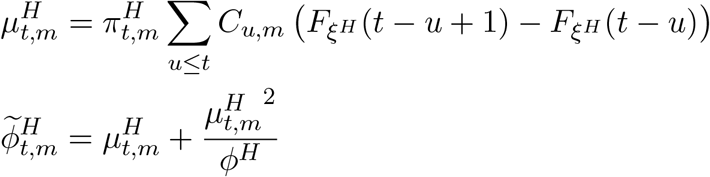

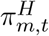 varies over time in the same way as 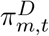to account for vaccination coverage and different variants. In contrast to 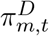, however, we can estimate 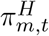 for each geographical region and do not have to consider it to be known. Doing so is important because medical care and definitions of hospital admissions and ICU admissions may vary between geographical regions. Therefore, we define a ratio 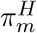 which is specific to each geographical region, but that does not change over time, and calculate 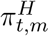 as a product of 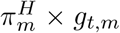 where *g*_*t,m*_ is a fixed quantity representing the effect of vaccinations and new variants that can potentially modify the severity of the disease.

We use exactly the same model for hospital (normal beds) and ICU occupancy. For the sake of brevity, we therefore do not present the model for ICU occupancy in detail, but it can be obtained by merely changing the superscripts from *H* to *Hicu*. The two distributions, 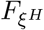 and 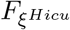, indicate the probability that a person with symptom onset on day *x* occupies a hospital or an ICU unit on day *x* + *y* with *y* = 1, 2, 3, For the application to COVID-19, we obtain these two distributions by combining information on the time between symptom onset and hospitalization with information on the time a person occupies a bed or ICU after being hospitalized through Monte Carlo methods. See sections C.3 and D.2 in the supplementary material for a more detailed description of the definition of 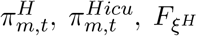 and 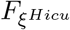.

We provide a summary of the model and the expression of the joint posterior in section A of the supplementary material.

## 3 Inference, identifiability and implementation

Due to the complexity of the hierarchical model, there is no analytical solution and we use a Metropolis-Hastings algorithm (Hastings, 1970) to sample from the joint posterior distribution. For the simulation study and the application, we fine-tune acceptance rates by using an adaptive phase (Brooks et al., 2011; Roberts and Rosenthal, 2009) and discard a defined number of iterations as burn-in. We apply thinning to reduce the autocorrelation in the generated Markov chains.

The flexibility of the proposed model can come at the cost of non-identifiability issues. The first obvious problem of identifiability occurs if we try to estimate the infection fatality rate 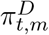, the probability of being hospitalized 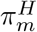 or being treated in ICU 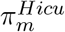 and the case detection ratios *ρ*_*t,m*_ simultaneously. This problem is easily circumvented by considering one of the four parameters as known. As the infection fatality rate can be reliably estimated in seroprevalence studies and modified by accounting for factors like the age structure of the population, vaccination coverage and the prevalence of different variants, we consider this factor known to be able to estimate the three remaining factors. The second identifiability issue arises in the estimation of the number of true and unknown infections. Since we assume that this variable follows a Negative Binomial distribution where the expected value is a function of the effects of non-pharmaceutical interventions (that are to be estimated), the model can in theory describe the data through any set of values for these parameters if the overdispersion parameter takes very high values. Moreover, the dimension of the latent variable can be rather high depending on the length of the observation window and number of geographical regions (the dimension in the latent variable grows with *t* and *m*) and it is not possible to use, for instance, Hamiltonian dynamics in the updating of this variable because it is not continuous. Current Bayesian hierarchical approaches commonly circumvent these problems by making the somewhat questionable assumption that the number of infections is a continuous variable that deterministically depends on the currently active NPIs (Flaxman et al., 2020; Brauner et al., 2021; Sharma et al., 2021). We address this issue by assuming an informative prior for the different overdispersion parameters in the renewal model, the hospitalization models and the death model and by splitting *I*_*t,m*_ in blocks of 10 for each geographical region *m* to be able to update each block one at a time. Finally, assuming a hierarchical model structure on *α*_*k,m*_, 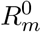 and *τ*_*m*_ has the advantage of stabilizing parameter estimates by using information across countries. This effect is particularly important for the estimation of non-pharmaceutical interventions. Since such interventions are often implemented or relaxed as multi-component interventions on the same or subsequent days in a country, it is difficult to disentangle their effects if we assume country-specific effects that do not follow a hierarchical structure because the estimated effects would be highly correlated. Using a hierarchical model allows us to account for variation in the effect of these interventions while using the information across countries to reduce the correlation between effect estimates. However, it is difficult to determine the exact amount of shrinkage that should be applied, expressed through the prior distributions on the variance parameters and this choice therefore needs to be transparently reported and tested in sensitivity analyses. We implement the algorithm in an object-oriented approach in Python 3 using the two scientific standard libraries `NumPy` (Harris et al., 2020) and `Scipy` (Virtanen et al., 2020) and accelerated computational critical parts with `Numba` (Lam et al., 2015) as a JIT compiler. The code is available on GitHub.

## 4 Simulation Study

### 4.1 Data generation

We carry out a simulation study with the aims 1) to assess the correctness of the implemented algorithm and 2) to investigate potential problems concerning the identifiability of model parameters. We generate 100 data sets with ten geographical regions and an observation period of 600 days for each of them. Thus each data set contains of 6,000 rows of data. We specify five artificial interventions with mean effects *α*_1_ = 0.22, *α*_2_ = 0.25, *α*_3_ = 0.3, *α*_4_ = 0.4, *α*_5_ = 0.45. This allows the basic reproduction number to be reduced to roughly 80% when all NPIs are active. To obtain region-specific effects of non-pharmaceutical interventions, we sample from a Gaussian distribution with the corresponding mean *α*_*k*_ and a standard deviation 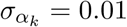. The basic reproduction number is sampled in the same way using a mean *R*^0^ = 3.25 and a standard deviation of 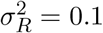. We seed the first day of the pandemic in each region by sampling from a Negative Binomial distribution with a mean *τ*_*m*_ which is generated from a Gaussian distribution with mean *τ* = 10 and *σ*_*τ*_ = 2. All overdispersion parameters of the Negative Binomial distributions are set to 1000 to obtain stable disease dynamics.

To obtain realistic time points at which the NPIs are set to active, we generate data in which the decision on whether a NPI is set to active depends on ICU occupancy: To do so, we generate Bernoulli variables for currently inactive NPIs at each *t* with probability *p*_*k,t*_ depending on ICU occupancy on *t* − 1. In the case a NPI is activated, it remains active for a random time period between 60 and 120 days.

The data generation is carried out in R version 4.0.4 (R Core Team, 2021). For further details, see the available R script that we used for the data generation.

### 4.2 Results on simulation data

We fit the model to each of the 100 data sets where we run 2 chains with 100,000 iterations and a burn-in of 10,000. We apply thinning by keeping only every 50th iteration. We check convergence by analyzing traceplots and potential scale reduction factors which are always *<* 1.01 Gelman and Rubin (1992).

As can be seen in Table 1, the algorithm produces estimates that are very close to the true NPI effects (with relative bias of at most 1.02%) and high coverage rates near to one.

**Table 1:**
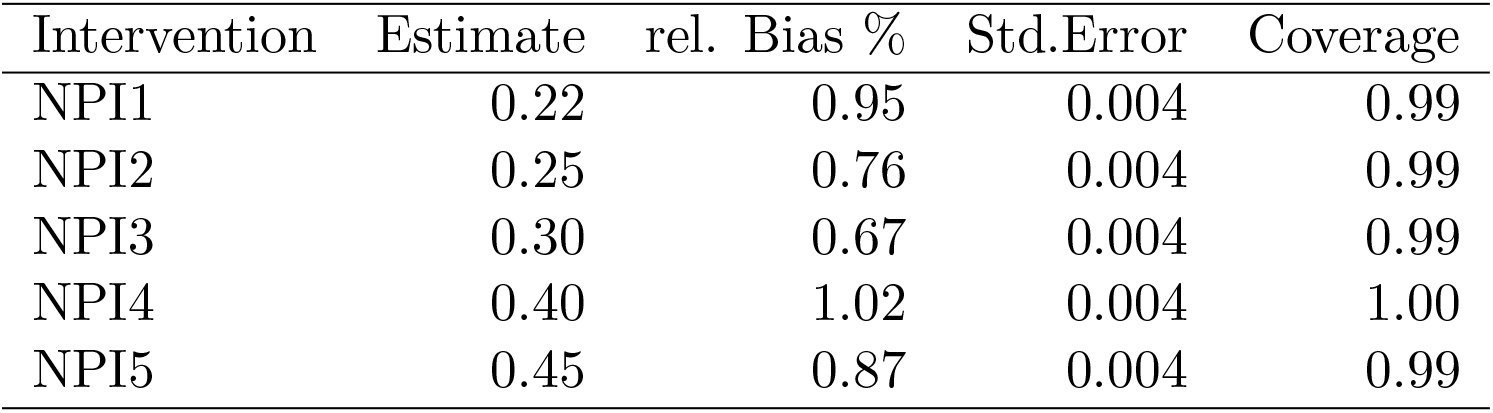
Average estimated effects of non-pharmaceutical interventions (NPIs) on simulated data. All values (except the coverage) are taken as the mean over all simulated datasets and generated Markov chains for all *α*_*k*_’s.

For illustration purposes, we present in Figure 2 the samples from the posterior as violin plot for the first three NPIs and 20 data sets. Figure 3 shows the posterior predictions of the number of (unknown) daily infections (3a) and reported cases (3b) for one of the ten regions for one data set.

**Figure 2:**
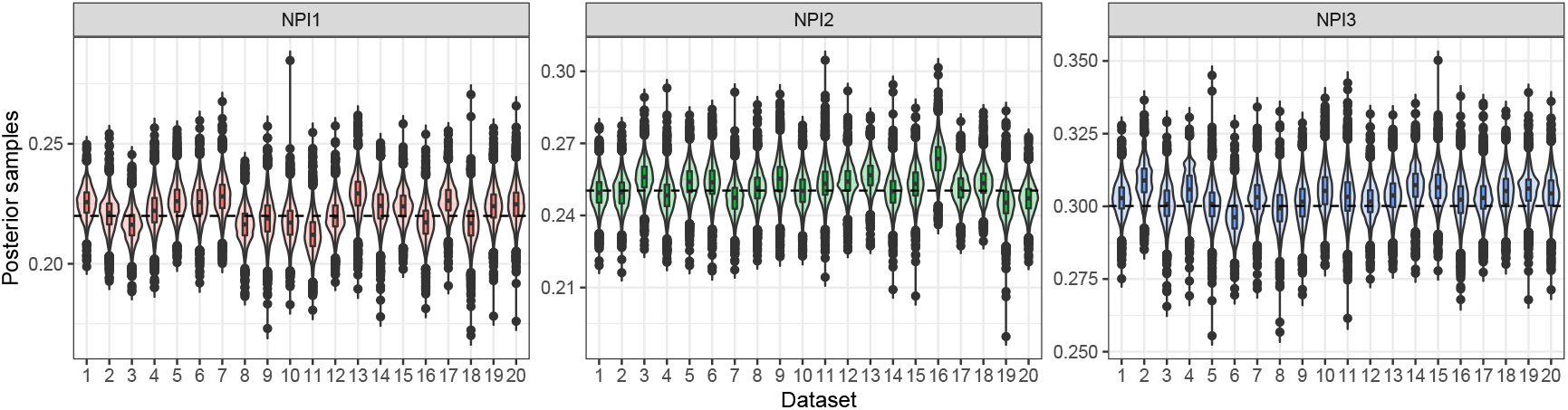
Results for the first three NPIs and 20 data sets. The horizontal line is the true mean value.

**Figure 3:**
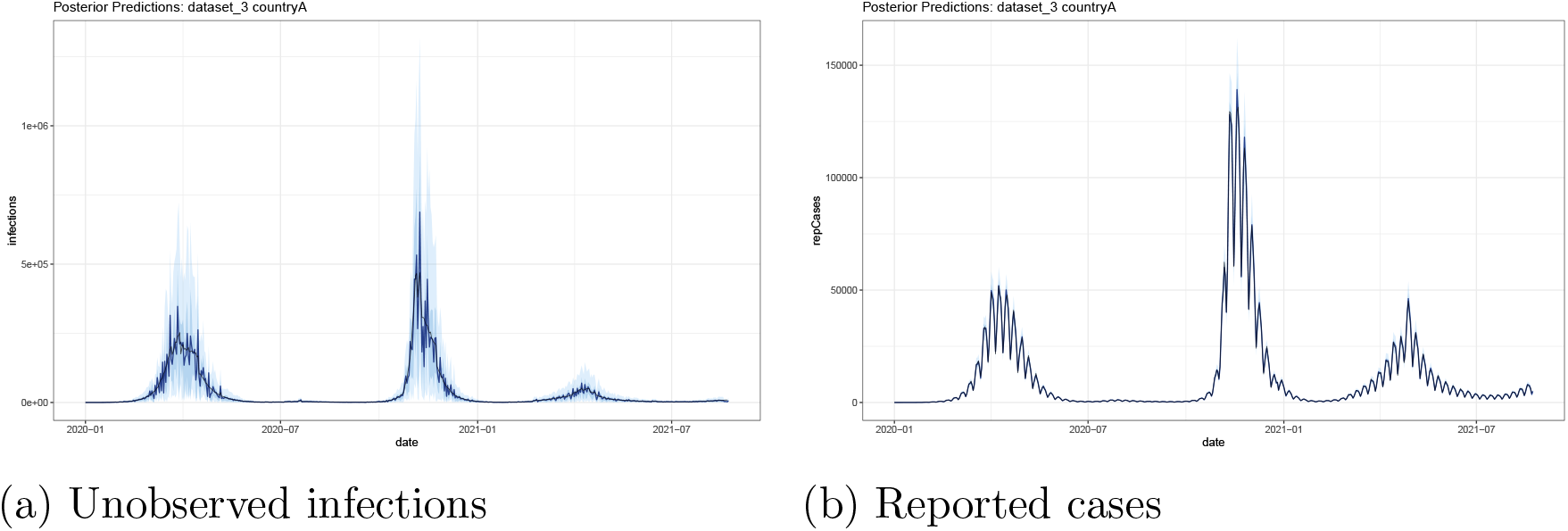
Posterior Predictions for two time series on simulated data. Black is the true underlying simulated time series. In blue the mean predictions with 95% credible interval.

## 5 Application

### 5.1 Data Sources

We apply our model to COVID-19 data of 20 European countries (Austria, Belgium, Czechia, Denmark, Finland, France, Germany, Greece, Hungary, Ireland, Italy, Netherlands, Norway, Poland, Portugal, Slovenia, Spain, Sweden, Switzerland, United Kingdom). Following Flaxman et al. (2020), we define the start of the observation period in each country as 30 days before ten cumulated deaths were reported. We include data on the whole course of the pandemic until the 31st of October 2021 resulting in a median length of 620 days. We define the following interventions using information from the COVID-19 Government Response Tracker (Hale et al., 2021) resulting in five NPIs: School closure, gatherings, lockdown, subsequent lockdown and general behavioral changes. This last NPI is active from the first time an NPI was implemented in a country and remains active until the end of the observation period. It subsumes many behavioral adaptations that were taken since the beginning of the pandemic and that remained more or less in place during the entire course of the pandemic. These might include less physical contact, working from home wherever possible and higher alertness in case of any respiratory disease symptoms, and wearing masks in some countries. More details on the definition of NPIs in our application can be found in section C.1 of the supplementary material. We use data on reported cases and deaths from the Johns Hopkins CSSE COVID-19 Dataset (Dong et al., 2020). Data on the prevalence of variants of concern and hospital and ICU occupancy are obtained from the *European Centre for Disease Prevention and Control* (ECDC) ^2^. Note that not all countries provide data on hospital occupancy (or only for part of the observed period). We address this issue by using only the terms of the renewal and the death model in the updating of the number of infections for countries for which there is no information on hospital and ICU occupancy. However, we can still produce posterior predictions for these countries by sampling among the current values of 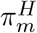 and 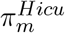 for an arbitrary country *m* at each iteration. Since the data on the prevalence of different variants is only available on a weekly basis, we fit a sigmoid function with a squared loss to obtain smooth daily data. More details on this procedure are presented in section C.2 of the supplementary material. The prevalence affects the model in the renewal equation and also in the IFR. Data on vaccinations are obtained from *Our World in Data* (Mathieu et al., 2021). Since we use a weighted IFR by age strata, we need information on the number of vaccinations in different age groups. However, very few countries provide information on the age structure of currently vaccinated individuals. We therefore use publicly available data from France and map the relative age-specific vaccination progress to other countries, making the assumption that the prioritization of vaccinations for different age groups evolved roughly in the same manner across different European countries. More details about the construction of the IFR can be found in section C.3 of the supplementary material.

### 5.2 Results

We run eight chains with a burn-in of 20,000 followed by 50,000 iterations per chain. We apply a thin of 100 resulting in 4,000 samples from the posterior distribution for each parameter. We run a longer adaptive phase with 200 adaptive steps (each with 100 iterations) to get good proposal standard deviations. For the final sampling procedure we again fine-tune these proposals by running ten adaptive phases (with 50 iterations each). Information about the convergence diagnostic for the parameters of major interest (NPIs and seasonal effects) are presented in the supplementary material E.5.

#### 5.2.1 Estimated effects of NPIs

Figure 4 and Table 2 provide information on the estimated reduction (obtained through 1−exp(−*α*_*k*_)) and increase (obtained through exp(−*α*_*k*_) −1) in the reproduction number for NPIs and seasons, respectively. For NPIs, the smallest effect is ‘school closure’ with a credibility interval that includes zero. The most effective NPI is ‘general behavioral changes’. When comparing the effects for ‘lockdown’ with ‘subsequent lockdown’, we can see that the first lockdown is estimated to have a larger effect than subsequent lockdowns, reflecting the fact that the first lockdown was characterized by stronger restrictions and probably better adherence to these than subsequent lockdowns. As expected, one can observe a strong seasonal influence with an estimated increase in the reproduction number of about 30% and 37% for autumn and winter, respectively.

**Table 2:** Mean and 95% intervals of posterior distribution for the mean effects and of predictive distributions for country-specific effects for NPIs and seasons. Note that the parameters are transformed by 1 − exp(−*α*_*k*_) and 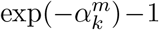to interpret them as relative reduction in percent for NPIs and as relative increase in percent for seasons.

| NPI | Mean | Pred |
| --- | --- | --- |
| School closure | 0.056 [-0.003, 0.111] | 0.057 [-0.132, 0.221] |
| General behavioral changes | 0.71 [0.694, 0.725] | 0.71 [0.662, 0.75] |
| Gatherings | 0.222 [0.176, 0.266] | 0.222 [0.076, 0.343] |
| Lockdown | 0.361 [0.283, 0.436] | 0.363 [0.17, 0.514] |
| Subsequent lockdown | 0.205 [0.141, 0.269] | 0.205 [0.041, 0.341] |
| Spring | 0.138 [-0.193, -0.084] | 0.137 [-0.276, -0.01] |
| Autumn | 0.303 [-0.354, -0.254] | 0.304 [-0.443, -0.177] |
| Winter | 0.373 [-0.445, -0.306] | 0.373 [-0.491, -0.269] |

**Figure 4:**
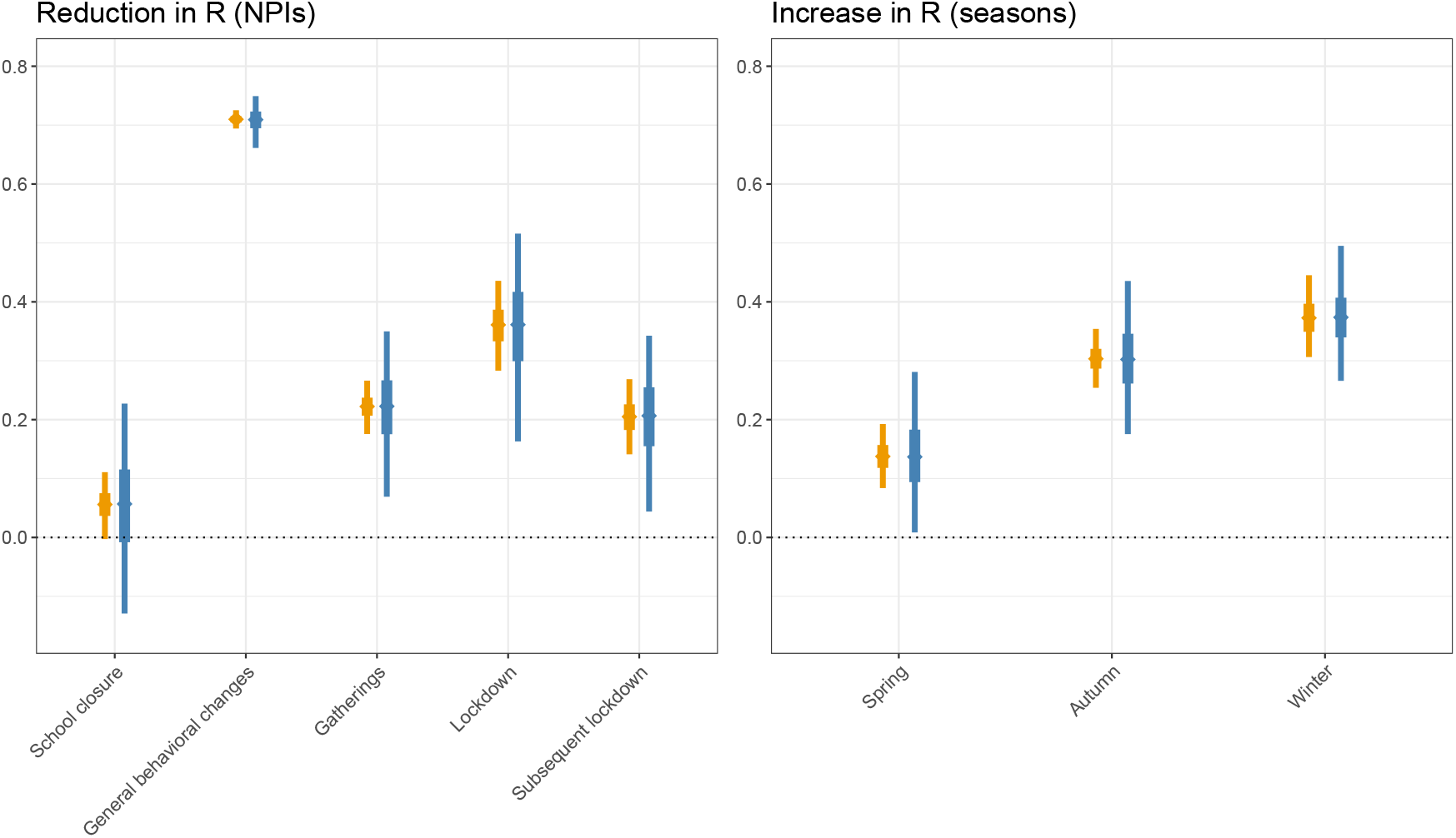
Estimated reduction and increase in the reproduction number for NPIs (left panel) and seasons (right panel), respectively. Posterior distributions for the mean effects *α*_*k*_ are given in orange. Posterior predictive distribution for *α*_*k,m*_ reflecting effect heterogeneity across countries are shown in blue. They are obtained by sampling from a normal distribution with mean *α*_*k*_ and standard deviation 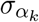 for each iteration. 50%- and 95%-credible intervals are given as bold and normal lines, respectively.

#### 5.2.2 Case detection ratios and posterior predictions

Figure 5 shows the estimated case detection ratios for two selected countries, Italy and France, for the entire observation period. The color encodes the estimated standard deviation, reflecting uncertainty in the estimation of these parameters. Furthermore, we provide information on the inverse test positivity rate in orange, i.e., the number of tests that must be performed to detect a positive case. In general, we observe high under-reporting (i.e., very small detection ratios) for the first wave of the pandemic indicating that the true number of infections by far exceeded the reported number of cases. Subsequently, the case detection ratios increase during the summer months and even reach values of up to 300%, i.e., there are three cases being reported for each true infection. This can be explained by the fact that the prevalence of the virus was very low during this period and the number of performed tests was very high. In this situation, there may be a non-negligible proportion of false positive results and we can therefore expect the number of reported cases to exceed the number of true infections due to the imperfect specificity of the tests (Brownstein and Chen, 2021; Cohen and Kessel, 2020; Kumleben et al., 2020; Bisoffi et al., 2020). Note, that these effects occur in situations where there are few prevalent cases, but a large number of tests are being performed. However, these results must be interpreted with caution as the estimated case detection ratios critically depend on the assumed value of the infection fatality rate and on assumptions about how this case fatality rate changes as a function of new variants and vaccination coverage.

**Figure 5:**
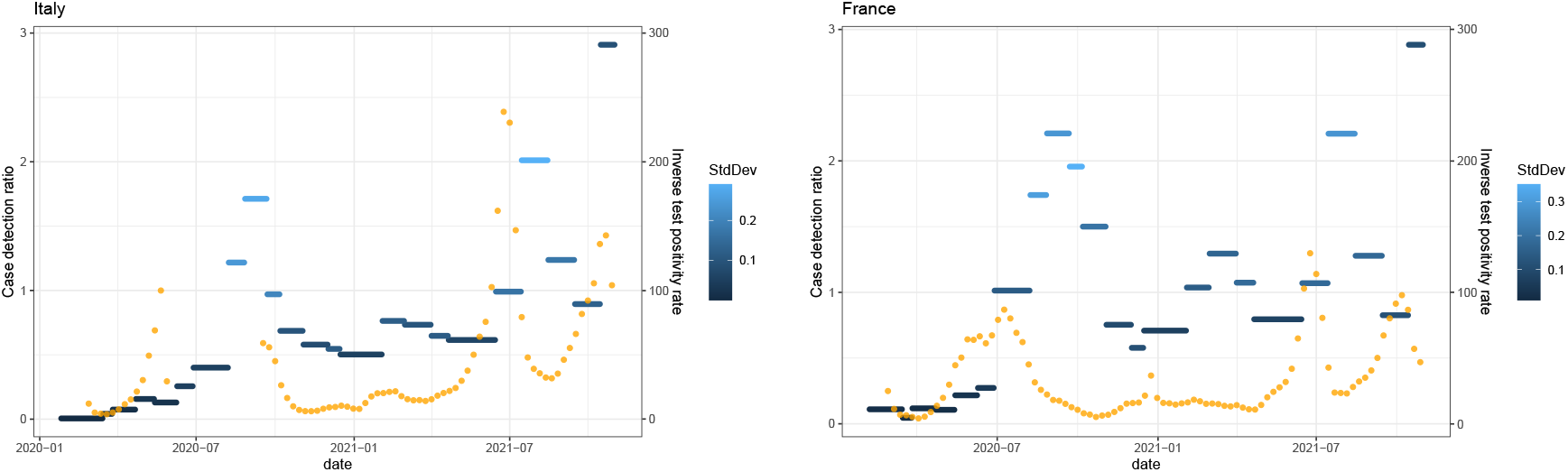
Estimated case detection ratios over the whole estimation window for Italy and France. The shades of blue represent the standard deviation of the posterior. In orange, we provide the inverse test positivity rate.

Figure 6 shows posterior predictions for reported cases, deaths and hospital occupancy for Italy and France. Black encodes the observed time series, the posterior mean and 50%- and 95% -credible intervals are given in blue. The approach captures the weekly variation in reported cases and deaths which are specific to the two countries. Moreover, it is capable of reproducing the three complementary time series, even though they provide quite contrasting information, in particular for the first wave. As we did not include data on hospital occupancy for Italy, the model uses the information on the proportion of hospitalized cases and estimated daily infections to predict this data. The posterior predictions and the case detection ratios for all countries are presented in sections E.3 and E.4 of the supplementary material.

**Figure 6:**
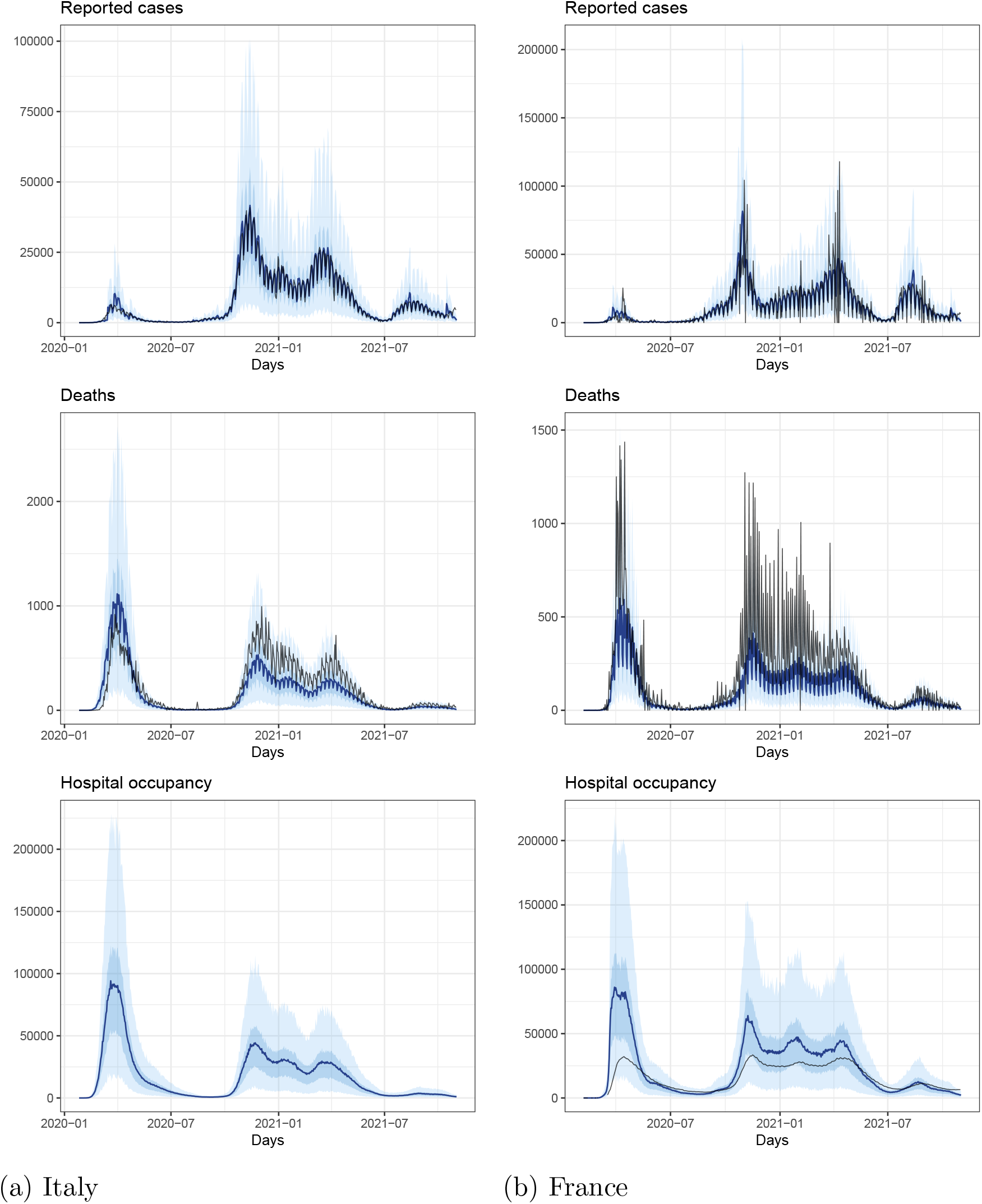
Posterior predictions of the reported cases for two countries. Black encodes the observed time series and blue the estimated mean with 50%- and 95% credible intervals

## 6 Discussion

In this work, we presented a Bayesian hierarchical approach to estimate the effects of NPIs. In this approach, we account for time-varying under-reporting, seasonality, the spread of the alpha and the delta variant and for vaccination coverage, allowing us to use the information on the entire course of the pandemic rather than focussing on short time periods during which the testing strategy remained constant. Owing to its modular nature, it is possible to use all available information while accounting for different sources of uncertainty in this information. Additionally, model and parameter assumptions can be transparently reported and the approach is very flexible, making it straightforward to adapt it to account for additional factors that might have an influence on disease dynamics.

Despite evidence on the importance of asymptomatic infections in the transmission of COVID-19, we do not explicitly distinguish symptomatic and asymptomatic cases. However, it is not clear whether this distinction would necessarily improve the model. Neither health registries nor seroprevalence studies systematically distinguish symptomatic and asymptomatic cases. As a consequence, it would be very difficult to ascertain which of the reported cases were symptomatic and to determine infection fatality rates that apply only to symptomatic cases.

In contrast to many other modelling approaches, we explicitly account for weekly patterns in the reporting delay distribution and it is therefore not necessary to smooth the time-series data on reported cases and deaths. As a consequence, we can model disease dynamics as a function of other influencing variables that may show variations on a daily scale. In future studies, it would be straightforward to integrate additional data, for instance on the number of performed tests, on weather conditions, from seroprevalence studies, or on measurements from wastewater.

In many countries, the question of whether public health measures should be implemented as a function of reported cases or hospital occupancy was widely debated as both quantities are to some extent unreliable. The proposed Bayesian hierarchical approach provides a framework in which information on both quantities (and on reported deaths and ICU occupancy) can be integrated to predict health care demand in the near future to be able to weigh costs, benefits and uncertainties in evidence-informed decision making.

## Data Availability

Links to public data are integrated in the code. All data produced in the study can be replicated with code from https://github.com/RaphaelRe/BayesModelCOVID

## A Summary and joint posterior distribution

Since the full model is a combination of many sub-models, we provide a summary in written form. We also provide the joint posterior for completeness.

The full model contains a latent variable *I*_*m,t*_ with dimension *T* × *M* (with *T* as the maximum number of used time points and *M* the number of geographical regions) where each *I*_*t,m*_ depends on the past {*t* − 1, …, 1} in each geographical region *m* and is seeded with a Negative Binomial distributed *I*_1,*m*_ where the mean is hierarchically given by *τ*_*m*_ with *τ*_*m*_ ∼ *N*^+^(*τ, σ*_*τ*_). This latent variable represents the true underlying number of infections which is unknown and assumed to follow a Negative Binomial distribution with overdispersion parameter *ϕ*^*I*^. We combine the information on the number of reported cases in the reporting model (with parameters 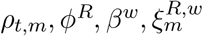), on the number of deaths in the death model (with parameters: 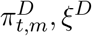) and possibly hospital occupancy and ICU occupancy in the hospitalization models (with parameters: 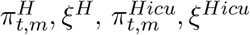) to estimate the number of true and unknown cases *C*_*t,m*_ with symptom onset at time *t*, which is a deterministic version of the number of infections *I*_*t,m*_, simply shifted with the incubation time distribution *ξ*^*C*^. Since there could be a strong weekly variation in the number of reported cases and reported deaths, we allow variations for certain weekdays for reporting delay *ξ*^*R*^ and the symptom-to-death distribution *ξ*^*D*^, respectively, through the parameters *β*^*R,w*^ and *β*^*D,w*^ where *w* indicates the weekdays (as part of 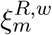 and 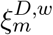). Given the latent variable, we estimate the effect of NPIs ***α*** in a hierarchical manner (controlled by the parameter 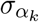), the basic reproduction number, also hierarchically (*R, σ*_*R*_) and the additional contagiousness of the new variants *β*^*alpha*^ and *β*^*delta*^. The vector of all parameters to be estimated is given by: 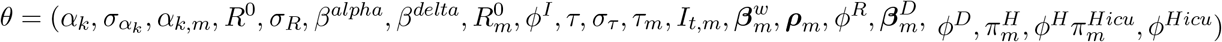 where a bold symbol denotes the collection of the parameters for brevity. The joint posterior distribution is given by:

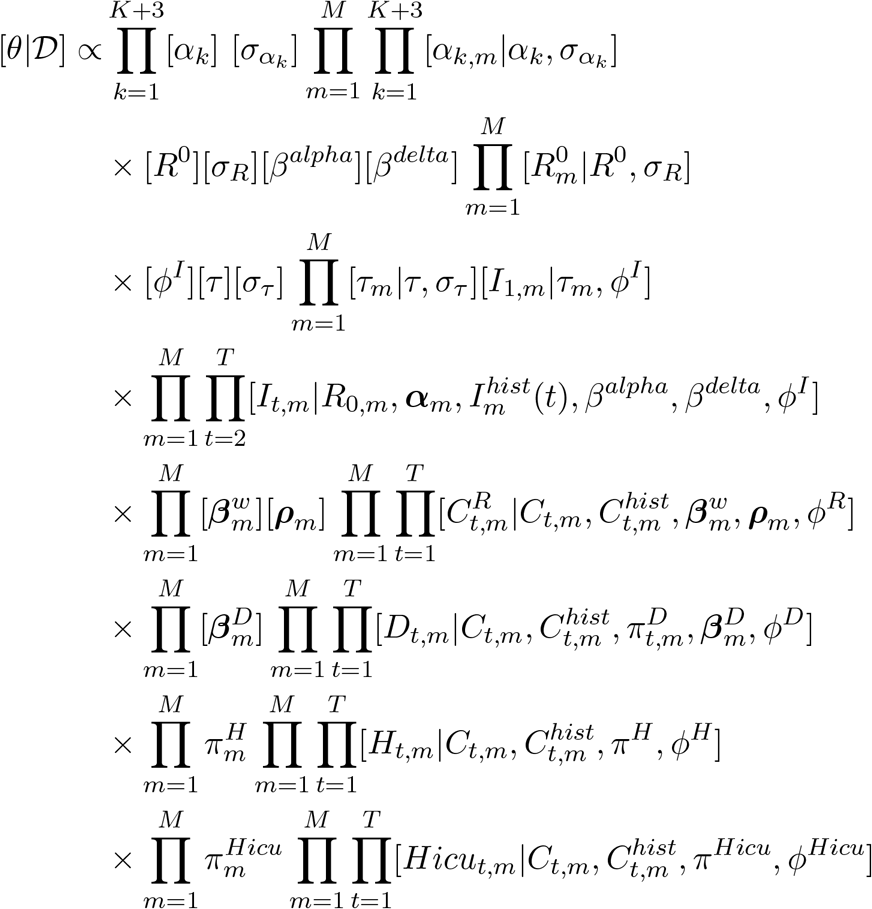

where 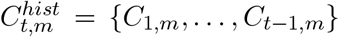 and 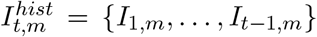. *C*_*t,m*_ is the shifted version of the infections *I*_*t,m*_ by *ξ*^*C*^. We write the conditional dependency in the death, reporting and hospitalization models in terms of the cases (*C*_*t,m*_) instead of the infections (*I*_*t,m*_) to highlight the time shift through the incubation time distribution.

## B Parameter and prior assumptions

### B.1 Time-shifting distributions

The model utilizes five different distributions (*ξ*^*I*^, *ξ*^*C*^, *ξ*^*R*^, *ξ*^*D*^, *ξ*^*H*^, *ξ*^*Hicu*^) to model the dynamics of the pandemic over time. Each of them is discretized to model daily frequencies. These discretized versions are used to shift a series in time while reflecting the uncertainty in the shifting time. The shift is done by a discrete convolution of the data with these distributions (see model description). Note that for *ξ*^*R*^ and *ξ*^*D*^ these are basic distributions which are getting modified by 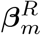 and 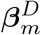 individually for each geographical region to model seasonality in the reporting. The bold symbol implies that the parameters are actually vectors, i.e., an atomic parameter to inflate or deflate the corresponding day of the week.

We specify the time-shifting distributions using information from Khalili et al. (2020), i.e., we take the reported means and the 95%-confidence intervals and choose the shape and scale parameters of gamma distributions in accordance with these quantities.

- *ξ*^*I*^: The generation time is assumed to be Gamma distributed with mean 5 and variation coefficient 0.45.
- *ξ*^*C*^: For the incubation time we assume a Gamma distribution with mean 5.68 and variation coefficient 0.08.
- *ξ*^*R*^: We estimate a basic reporting delay distribution empirically from data provided by the state office of health of Bavaria in Germany. The database offers detailed information about the individual history of patients. It is therefore possible to infer the time until reporting for a given weekday. More details can be found in D.1.
- *ξ*^*D*^: We model the time from symptoms to deaths as a Gamma distribution with mean 15.93 and variation coefficient 0.1.
- *ξ*^*H*^ / *ξ*^*Hicu*^: We model the hospital occupancy as a combination of the time from onset of symptoms to hospital admission and the time a person occupies a bed. The time from onset of symptoms to hospital admission is modeled as a Gamma distribution with mean 4.92 and variation coefficient 0.11. The time of occupancy is estimated from the bed allocations of Klinikum Großhadern, Munich. Details can be found in section D.2.

### B.2 Other parameters

For *β*^*vacc*1^ and *β*^*vacc*2^ we chose 0.6 and 0.3 respectively to reflect the heterogeneity in the used vaccinations and uncertainty over the actual effect (Pritchard et al., 2021; Voysey et al., 2021; Keehner et al., 2021; Polack et al., 2020; Baden et al., 2021). This results in an overall effectiveness of 90% after two vaccinations.

The probability of reinfection *β*^*reinf*^ is set to 0.16 (Hall et al., 2021).

### B.3 Prior distributions

We specify for the set of parameters 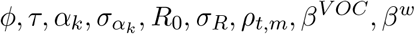 and *π*^*H*^, prior distributions as follows where Normal distributions (possibly truncated at zero) are always parameterized with mean and standard deviation, Gamma distributions with shape and scale and Uniform distributions with its minimal and maximal value respectively:

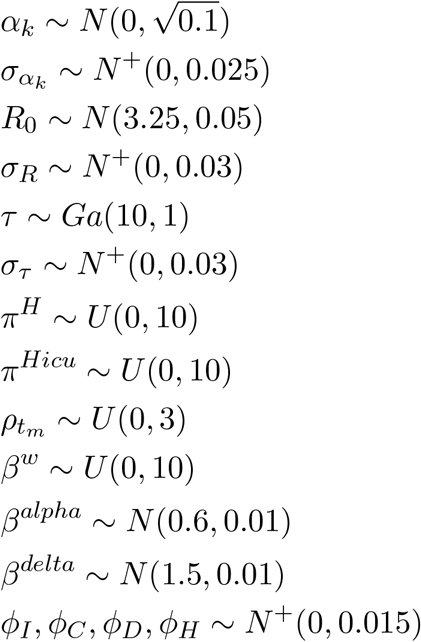

## C Data and Preprocessing

### C.1 Definition of NPIs

The definition of the NPIs is based on the COVID 19 Government Response Tracker (Hale et al., 2021). We take a subset of the provided time series and define the NPIs as follows:

- School closure: NPI is active when closing is required for at least some levels and a general targeting
- Gatherings: NPI is active when gatherings are restiricted to 10 or less people and a general targeting
- Lockdown: NPI is active when leaving house is prohibited (with possible exceptions such as daily exercise, grocery shopping, and ‘essential’ trips)
- Subsequent Lockdown: Since subsequent lockdowns often followed a much more detailed protocol we decided to encode them as separate NPI
- General behavioral changes: With the outbreak of the pandemic, people changed their general behavior (even in phases of low incidence). Therefore we define this NPI as active all the time starting when the first NPI gets active.

Figure 7 shows the NPIs as timeline for all countries. Note that some countries never implemented some of the NPIs. This is mainly the case for lockdown. Therefore there was no subsequent lockdown as well.

**Figure 7:**
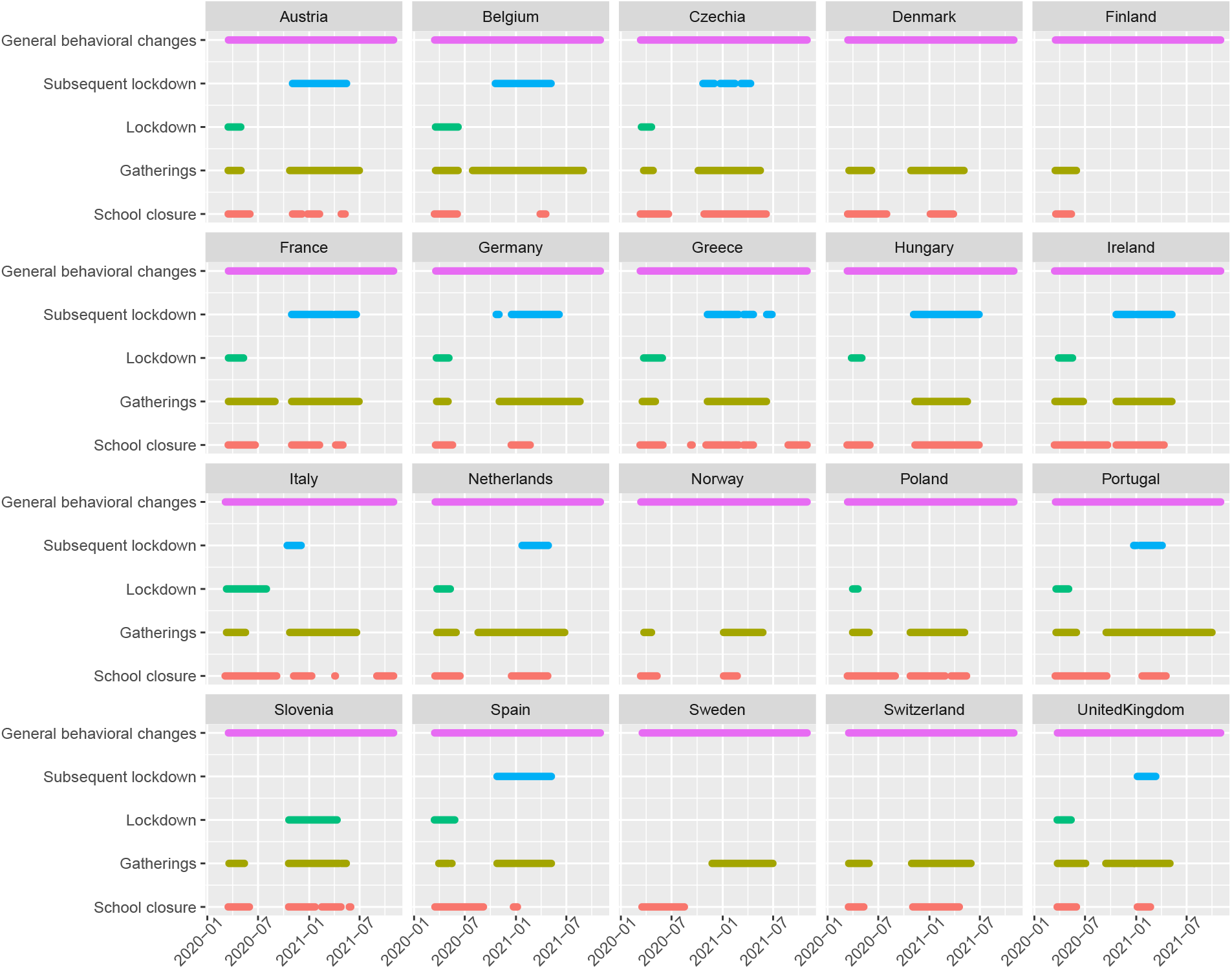
Timeline of the defined NPIs in all countries.

### C.2 Variants of Concern

To obtain the daily prevalence of the variants of concern, we fit a sigmoid function to the weekly data by minimizing the mean squared error. In this sense, the optimization problem is given as

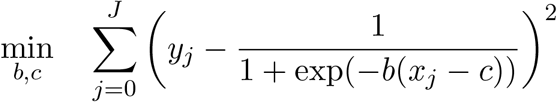

We use a *BGFS* algorithm as optimizer and run the optimization with 10 randomly sampled start configurations to get a good fit to the data.

The start of the used data window is seven days before the prevalence was first measured with a value *>* 0. End is the maximum of all provided prevalence data points in the country.

This procedure is independently done for the two variants. To find a good balance between the wild type and the two considered variants, we further use the following approximation to calculate the three proportions to get a value of one for all the variants. If 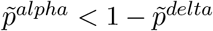 we set 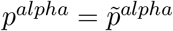. Else we use 1 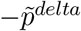. Where 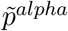 is the estimated prevalence from the fitted sigmoid function. *p*^*orig*^ is calculated as 1 − *p*^*alpha*^ − *p*^*delta*^ and 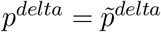 where again 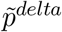 is the estimate from the sigmoid function.

### C.3 Infection Fatality Ratio

The infection fatality rate (IFR) gets affected by two quantities:

- Vaccinations: The probability to die after getting vaccinated is reduced drastically.
- Variants of concern: The new variants may have a much higher severity resulting in a much higher IFR.

Since the chance of a severe progression of the virus depends heavily on the age of an individual, we calculate a basic IFR as a weighted average between different age strata. Afterwards, we modify this basic IFR resulting in a time dependent IFR.

#### Weighted IFR by Age

We use information about the age strata of each country from O’Driscoll et al. (2021) and the aggregated IFR for four different age groups *g* from Staerk et al. (2021):

| Age group [in years] | 0-34 | 35-59 | 60-79 | 80+ |
| --- | --- | --- | --- | --- |
| $ifr_g$ [in %] | 0.008 | 0.122 | 0.992 | 7.274 |

Therefore the IFR in each country is defined as

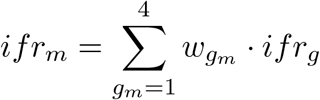

where 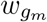 is the proportion of the age category of the population of country *m*.

#### Effect of vaccination

We include the effect of vaccinations by reducing the IFR in each age strata relative to their share of the population. Since most countries do not provide enough information about their vaccination progress in the different age groups, we choose to use publicly available data from Santé Publique France (2021) and adapt them to all other countries. We extract the relative proportions of people vaccinated in each age strata and map this proportion to the age strata of the other countries. This is a rather crude approximation, but unfortunately few countries provide data about their vaccinations on such a granular level. Moreover, the majority of European countries started by vaccinating vulnerable groups first, making it plausible to assume that the evolution of vaccination coverage over time in the different age groups was roughly comparable across different countries. We assume that after the first vaccination, the probability of dying is reduced by 80% after a lag of two weeks. For example, Haas et al. (2021) found a higher effectiveness against COVID-19-related deaths of the BNT162b2 vaccine. However, since not all countries use the same vaccinations and one can assume a reduction of the vaccine effectiveness as time goes on, we decide to use this crude approximation with a lower effectiveness.

#### Effect of Variants of Concern

Fisman and Tuite (2021) provide information about the severity of the variants of concern. We use our approximated time series for the prevalence (see section C.2 for details) and combine them with the information about the severity. This inflates the IFR and therefore leads to a higher number of deaths with an increasing proportion of the variants of concern when fixing the number of infections. For B.1.1.7 we inflate the IFR with 151% and for B.1.617 with 208%.

For more details of all steps described here, we refer to the file ‘calculate_weighted_ifr.R’ which is part of the provided code.

#### Adaption for 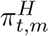

Since 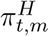depends on *t* and *m*, we also have to include the same effects forthe hospitalization which we formulate in the model as 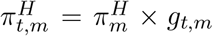where *g*_*t,m*_ is the effect of vaccinations and the variants of concern. Weapproximate *g*_*t,m*_ by calculating 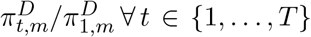. Therefore the model actually estimates an overall 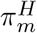 for each country separately and uses the transformed version 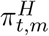 in the hospitalization models.

## D Estimation of *ξ*^*R*^, *ξ*^*H*^ and *ξ*^*Hicu*^

*ξ*^*R*^, *ξ*^*H*^, and *ξ*^*Hicu*^ are estimated from data and are not modeled as gamma distributions. Here we describe the procedure.

### D.1 Estimated reporting delay distributions for specific week-days

For the estimation of the reporting delay distribution, we use the COVID-19 reporting data of the Bavarian State Office for Health and Food Safety (LGL) which provides partial information about disease onset and time of reporting. We use this information to estimate the reporting delay in the data. We estimate seven discrete empirical reporting delay distributions, one for each weekday separately. We use a cut-off of 60 days and assume that a longer reporting delay does not exist. Figure 10 shows the estimated weekday-specific reporting delay distributions. For each day, there is a clear ‘dip’, i.e., one can observe a clear decline in the reporting on the 7th day for the estimation for Mondays, on the 6th day for Tuesdays, etc. The majority of the cases are reported on Wednesdays and Thursdays with around 15% to 20% each.

**Figure 8:**
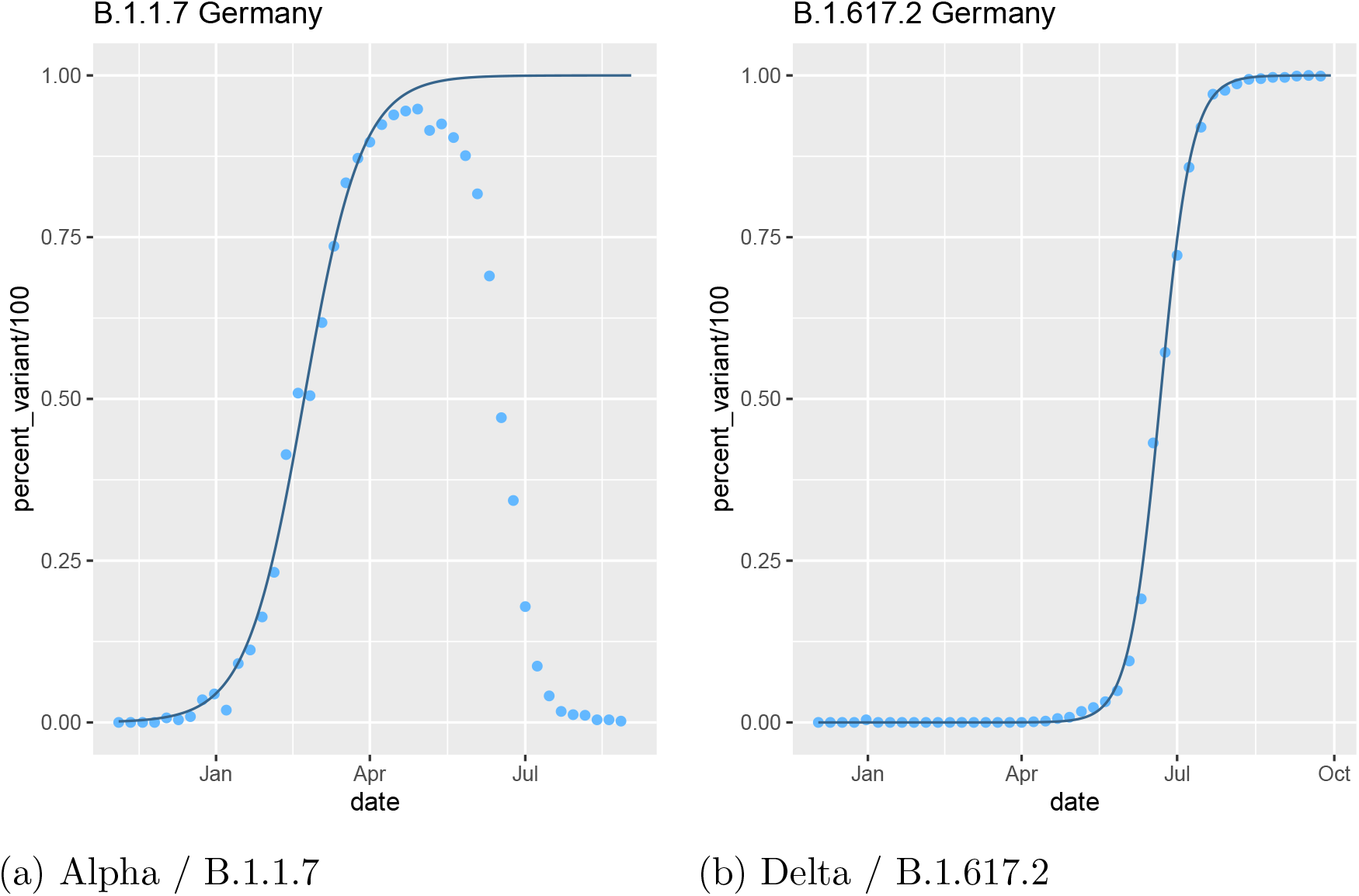
Estimated transition for variants of concern in Germany. The points depict the weekly data and the line, the fitted smooth curve.

**Figure 9:**
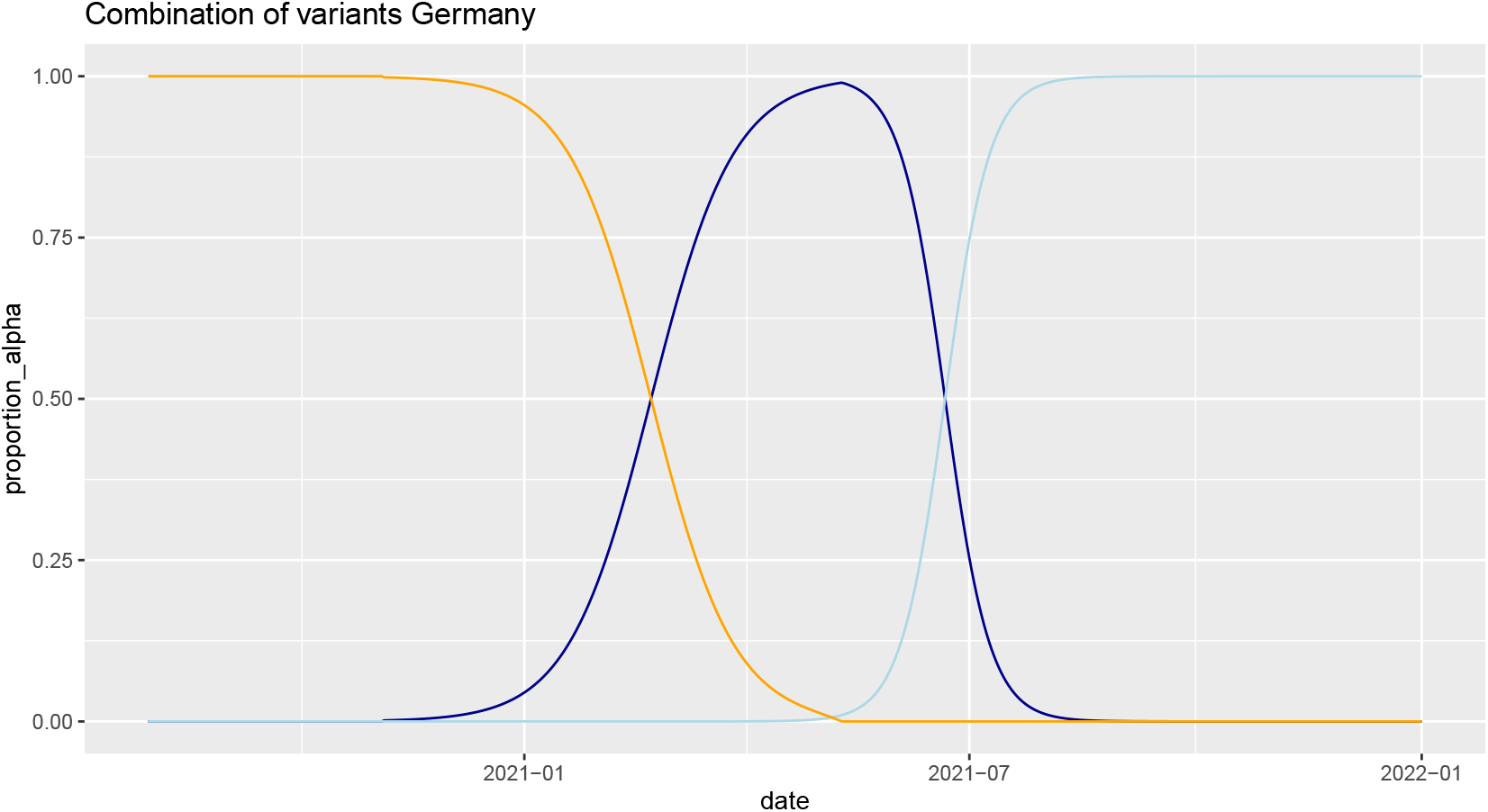
Combination of the three included variants. Yellow: original wild type, Darkblue: Alpha, Lightblue: Delta

**Figure 10:**
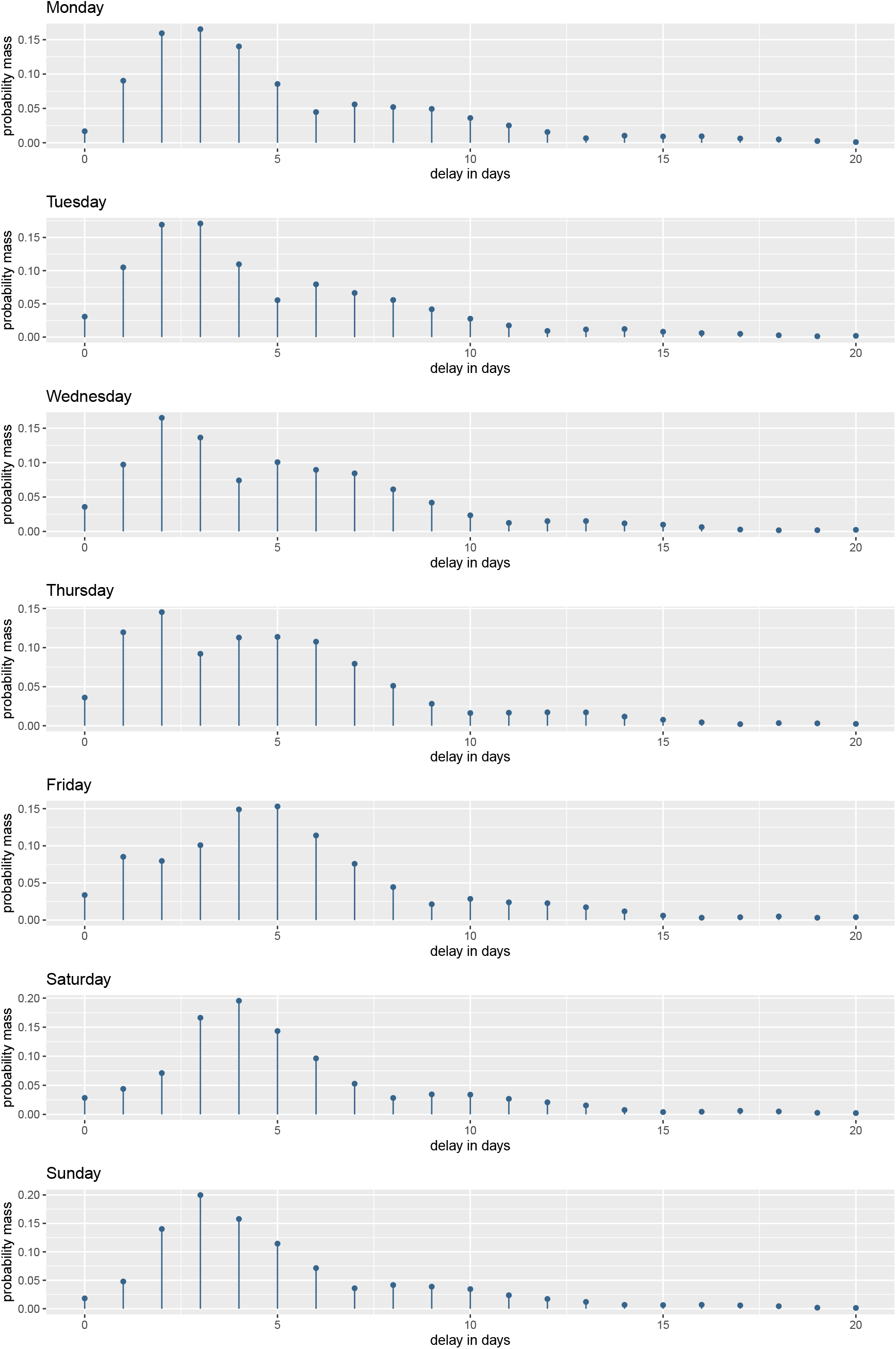
Estimated weekday-specific reporting delay.

### D.2 Estimation of *ξ*^*H*^ and *ξ*^*Hicu*^

To model the occupation of hospital and ICU beds we utilize *ξ*^*H*^ and *ξ*^*Hicu*^. Here we show how we calculate these quantities.

We combine the time from onset of symptoms to hospital admission and the time a person occupies a bed. The time from onset of symptoms to hospital admission is modeled as a Gamma distribution with mean 4.92 and variation coefficient 0.11. The time of occupancy is estimated on the bed allocations of Klinikum Großhadern Munich, Germany. For each person which got admitted to the hospital, first we sample a random time from the gamma distribution and then track the time the person lies in a normal bed or ICU unit. After doing this for all admitted patients, we sum the time of occupied beds for each day and re-normalize it by dividing with the overall sum resulting in an overall time distribution.

## E Results application

### E.1 Posteriors NPIs individual countries

Figure 11 presents the estimated posteriors for all countries individually. Note that not all NPIs were implemented in all countries. Therefore some, not all, *α*_*k,m*_ were estimated.

**Figure 11:**
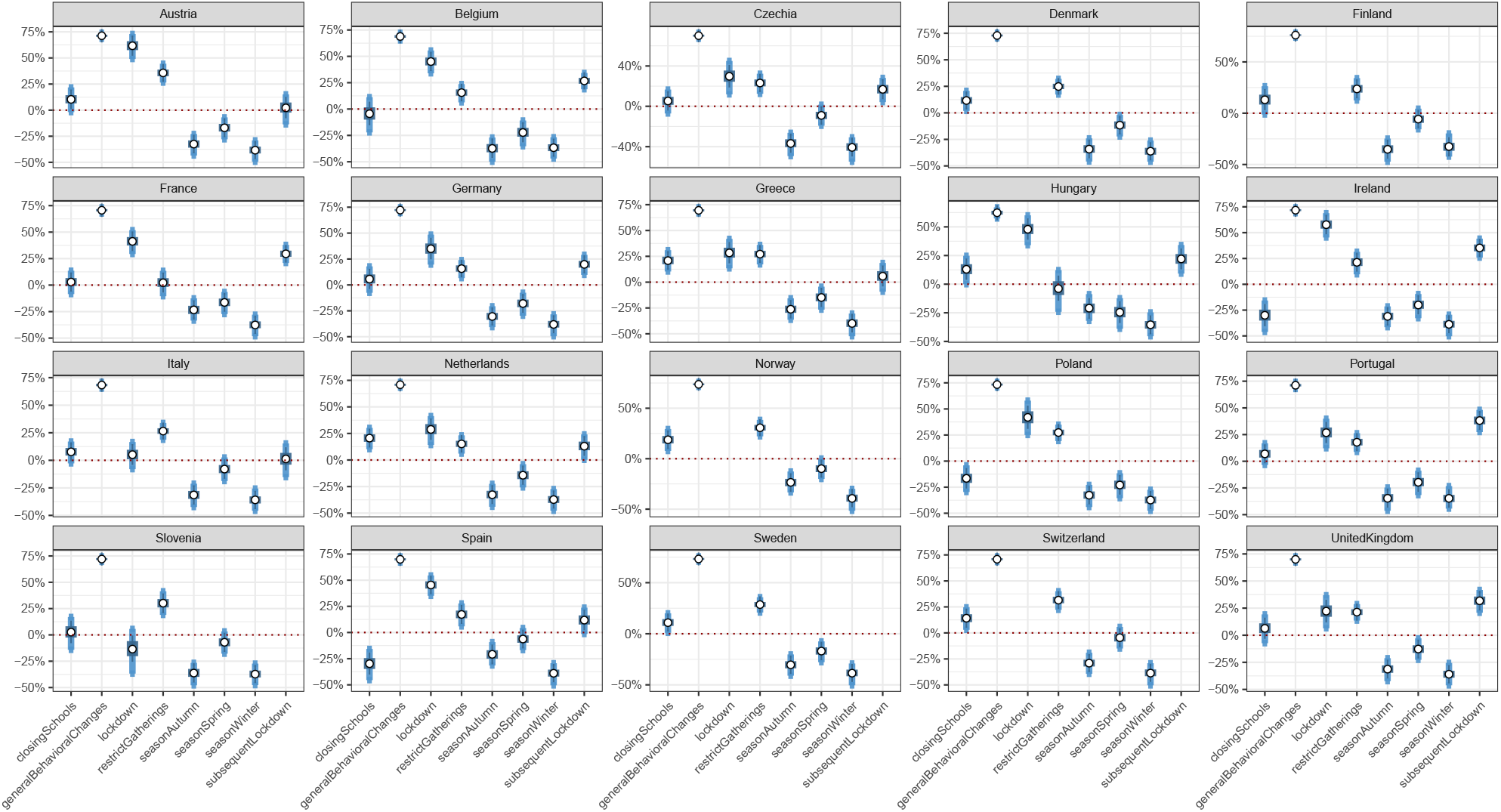
Mean, 50%- and 95%-credible intervals for all individual *α*_*k,m*_.

### E.2 Posteriors variants of concern

Figure 12 shows the prior and posterior distributions for the over-contagiousness of the variants of concern, i.e., *β*^*alpha*^ and *β*^*delta*^.

**Figure 12:**
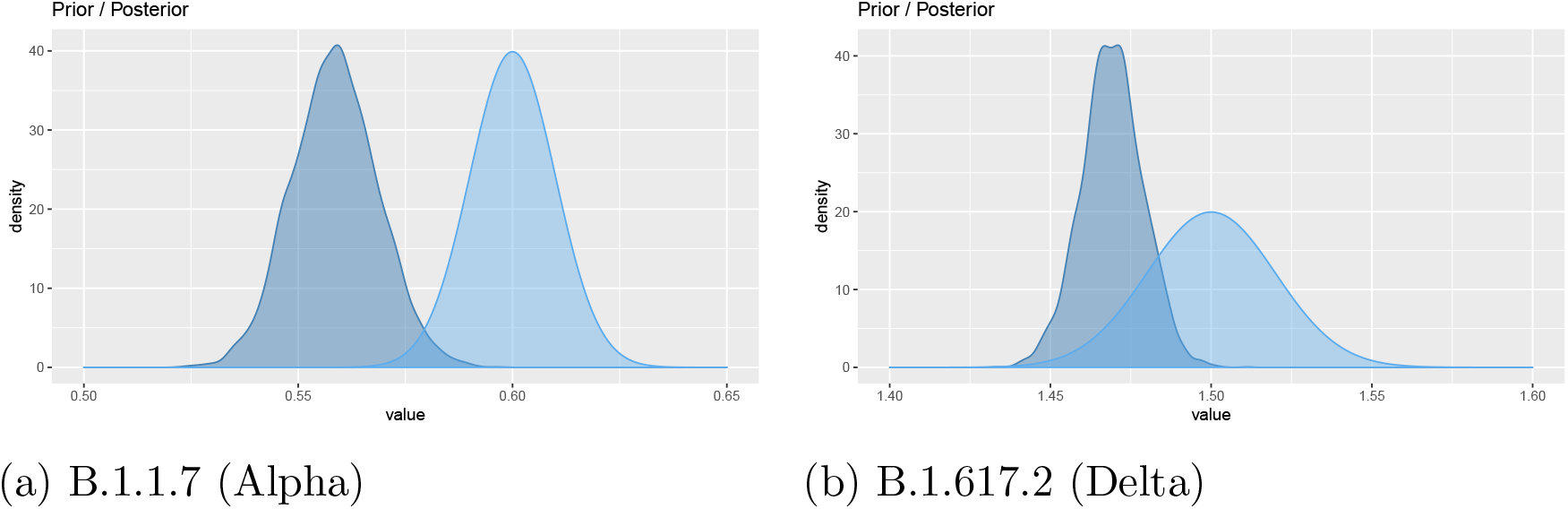
Prior (lightblue) and posterior (darkblue) for over-contagiousness of the variants of concern.

### E.3 Posteriors Predictions

Figures 13, 14, 15, 16 and 17 show the posterior predictions for all countries.

**Figure 13:**
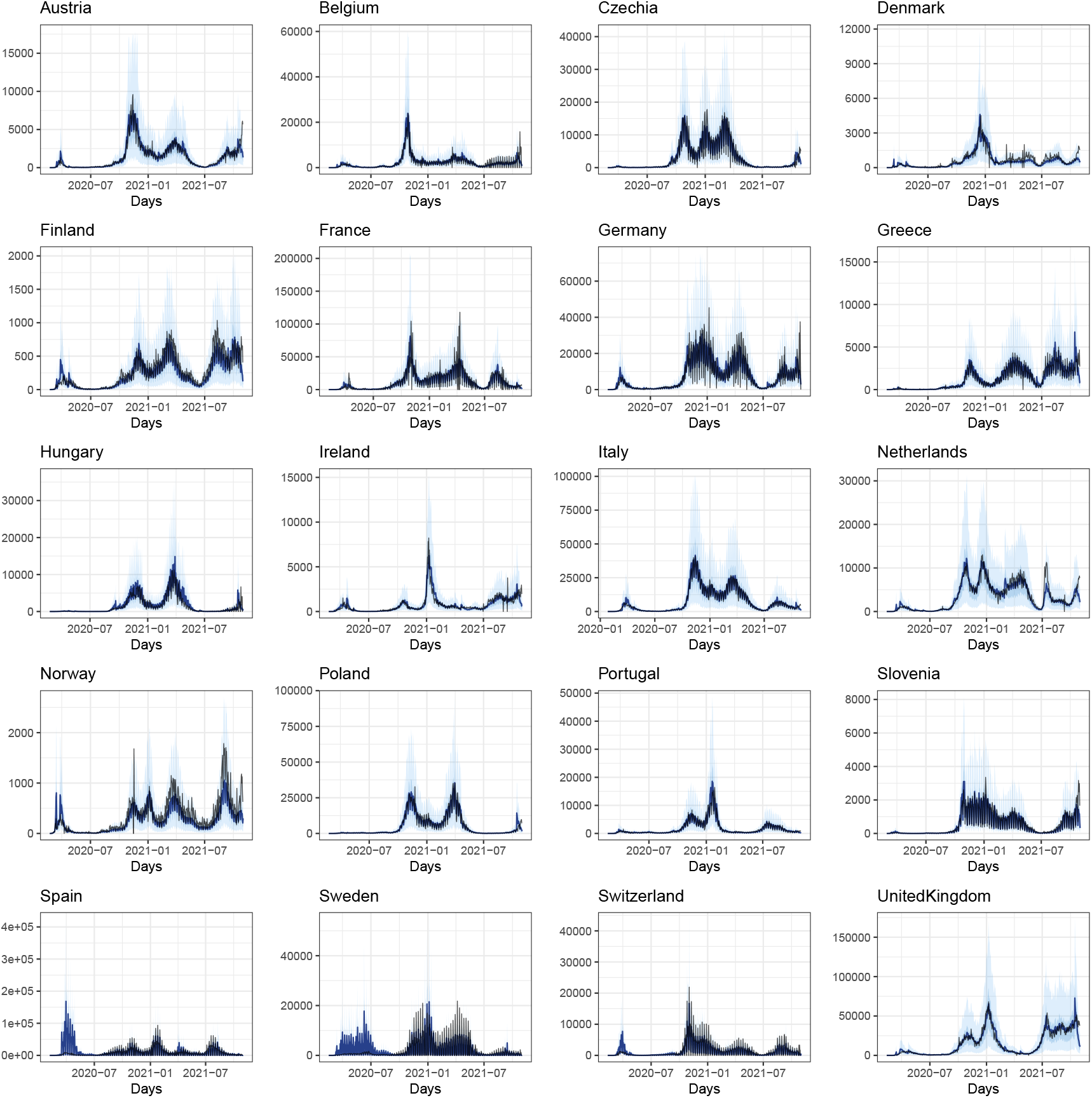
Posteriors predictions for reported cases

**Figure 14:**
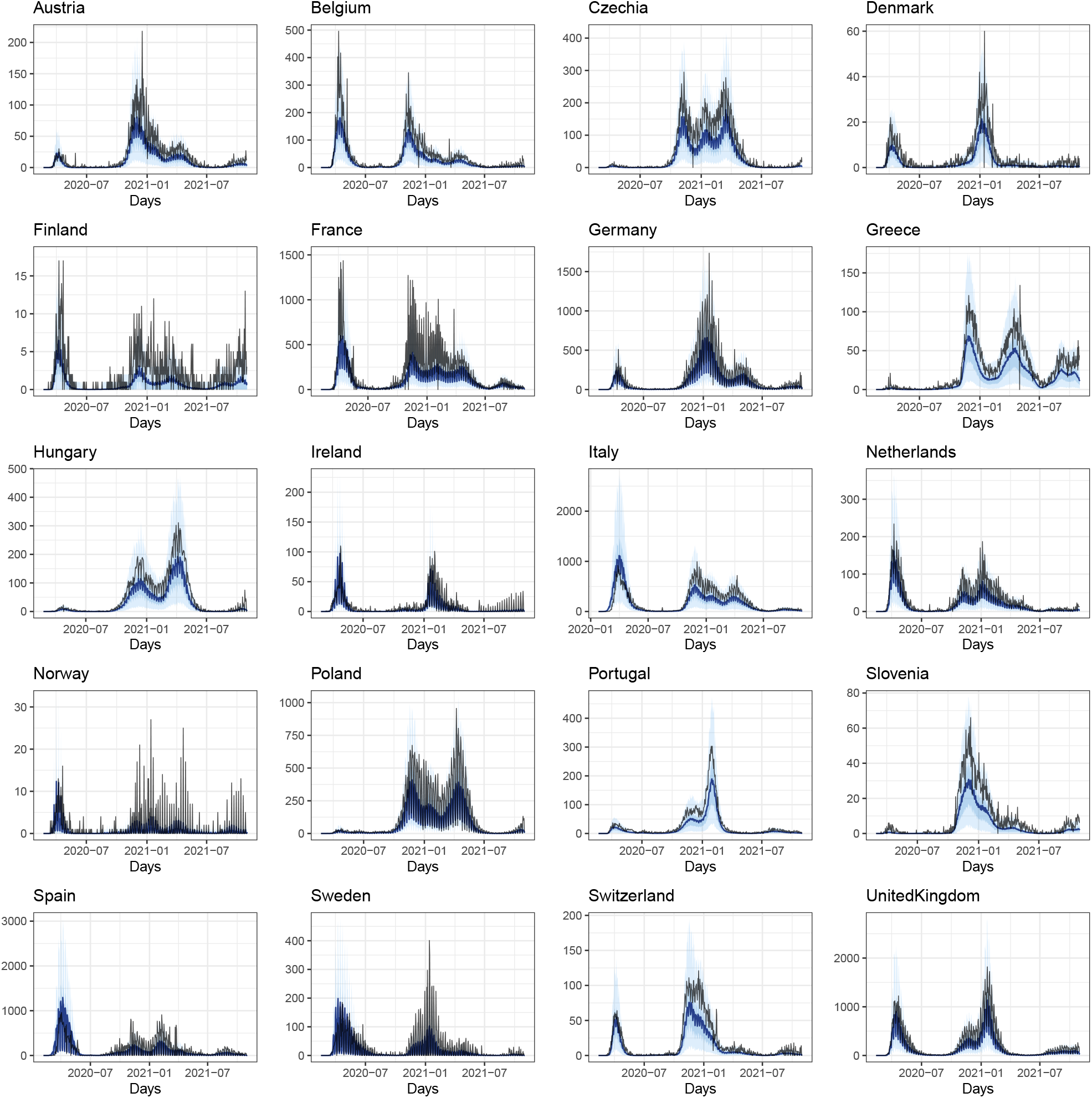
Posteriors predictions for reported deaths

**Figure 15:**
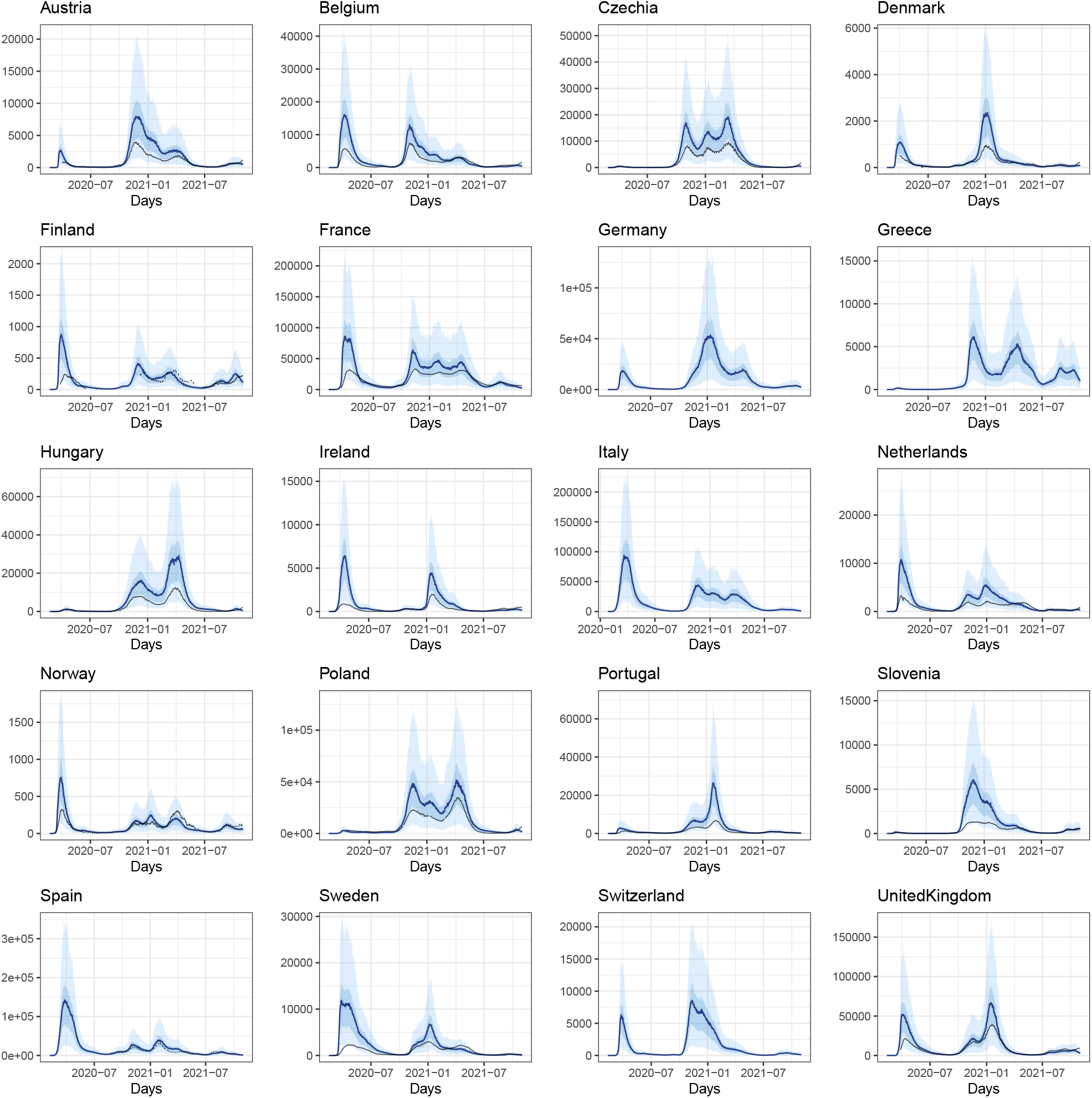
Posteriors predictions for hospital occupancy

**Figure 16:**
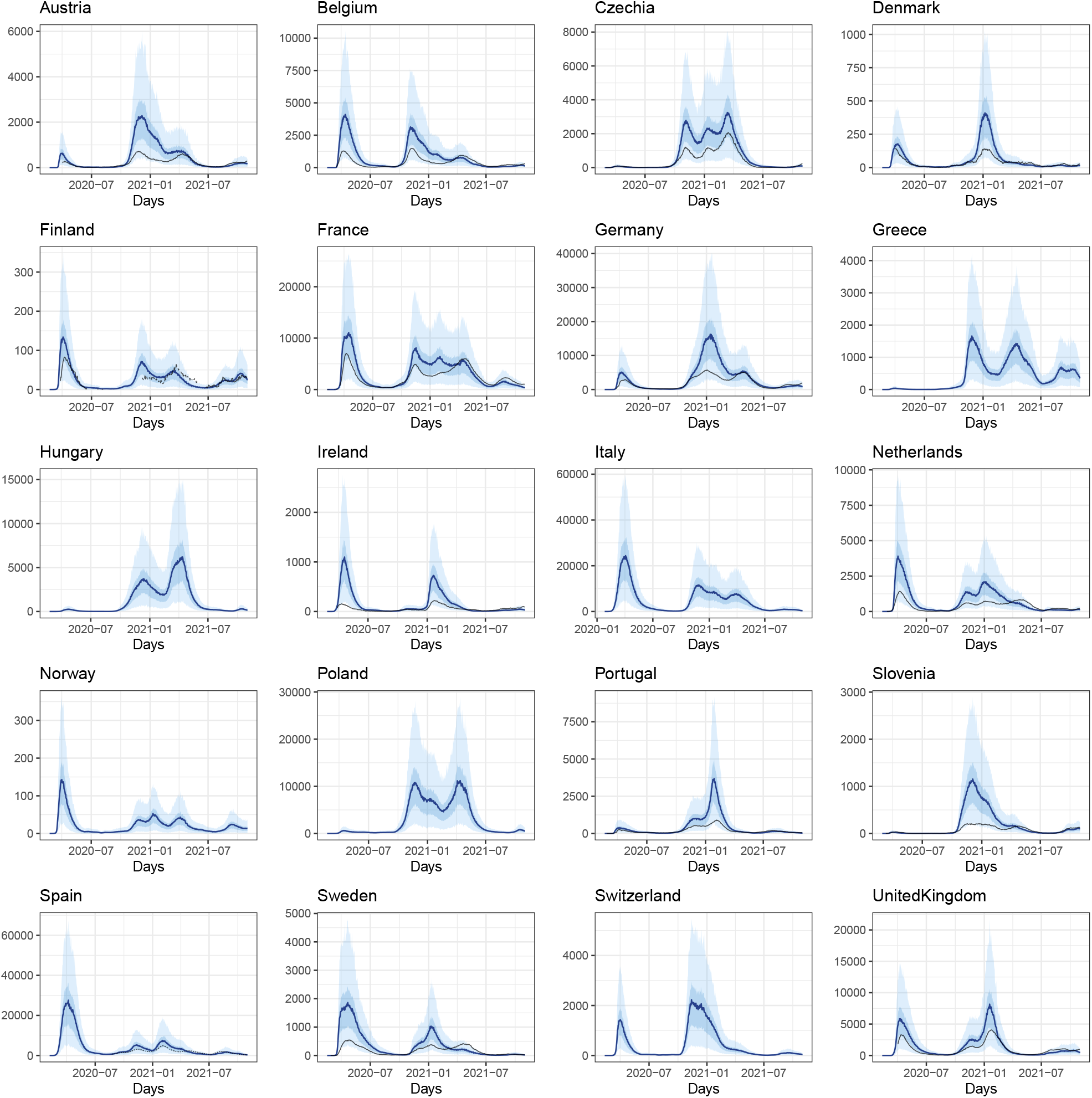
Posteriors predictions for ICU occupancy

**Figure 17:**
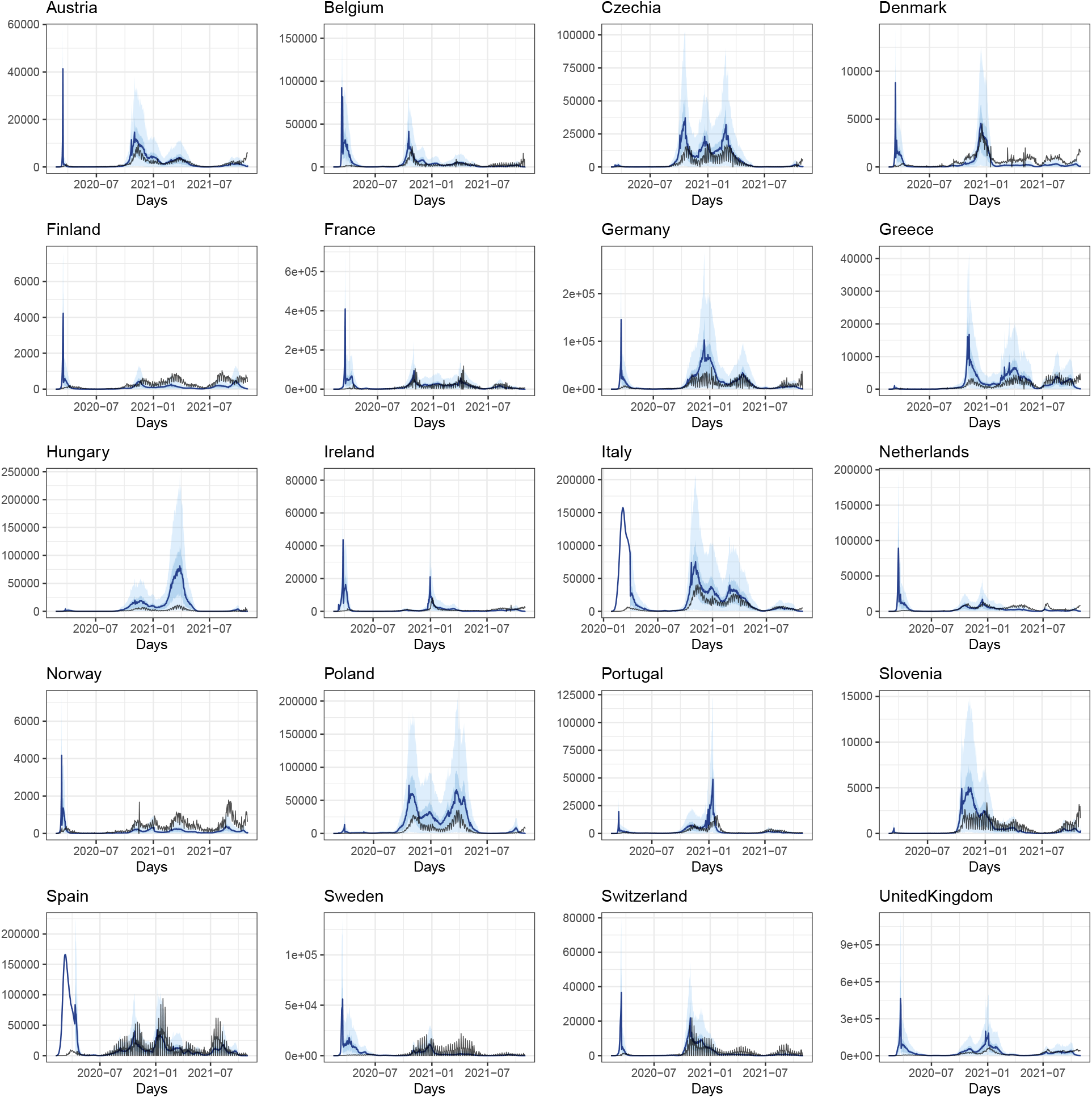
Estimated number of daily infections (in blue) and observed number of reported cases (in black)

### E.4 Case detection ratios

Figure 18 shows estimated case detection ratios for all countries. For some of the Nordic countries, the case detection ratio stays high indicating extensive testing over the whole observation period. The first period is sometimes estimated rather high where it would be expected to be low. However, here we observe a rather high uncertainty.

**Figure 18:**
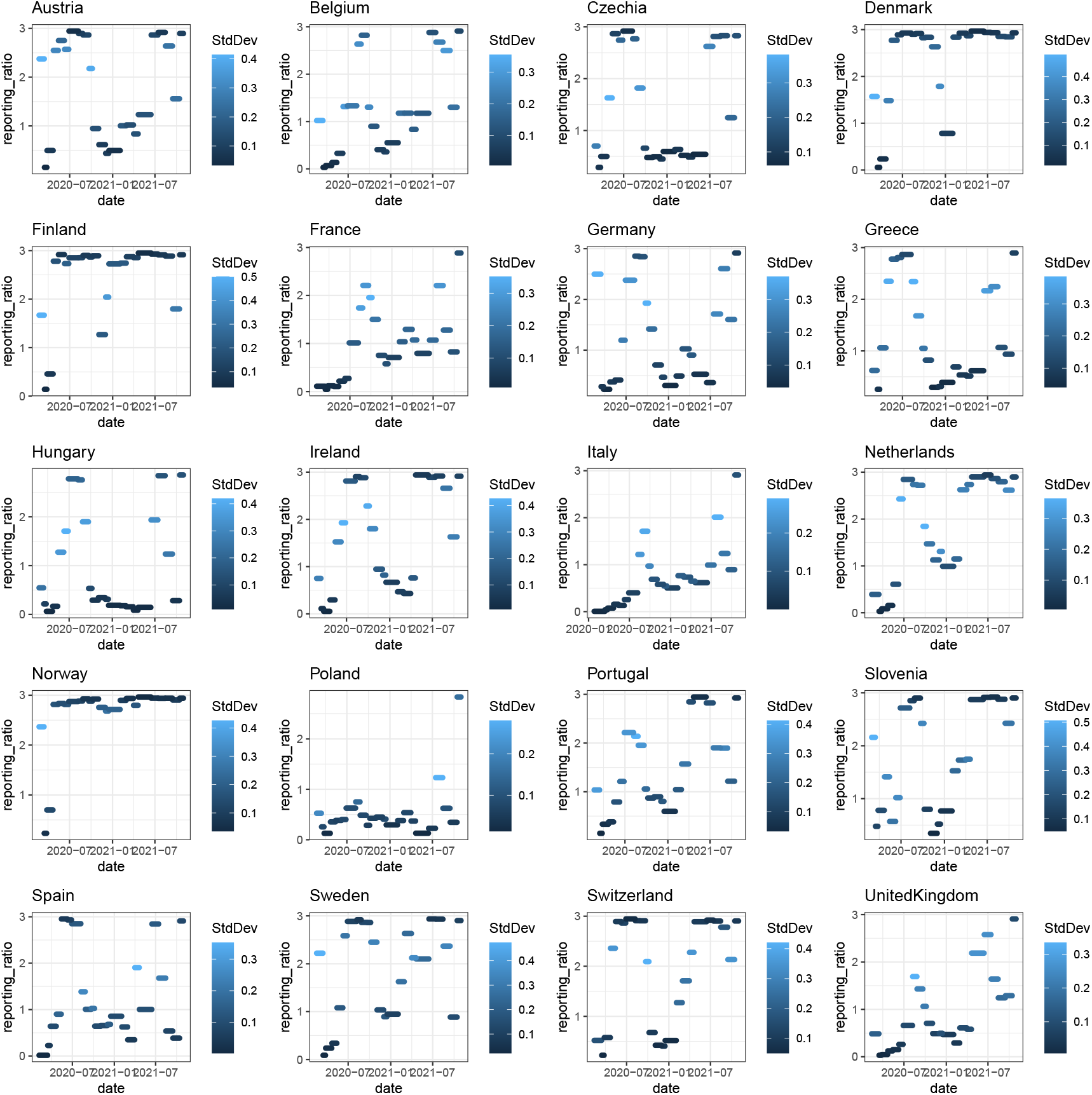
Estimated case detection ratios for *ρ* for all countries

### E.5 Trace Plots and convergence diagnostics

Figure 19 shows the trace plots of the mean effects of the NPIs and seasons. We observe sufficient values for the potential scale reduction factor between 1 and 1.05 (Gelman and Rubin, 1992). To obtain 4000 high quality samples, eight chains were used where each of them contains 500 samples after thinning.

**Figure 19:**
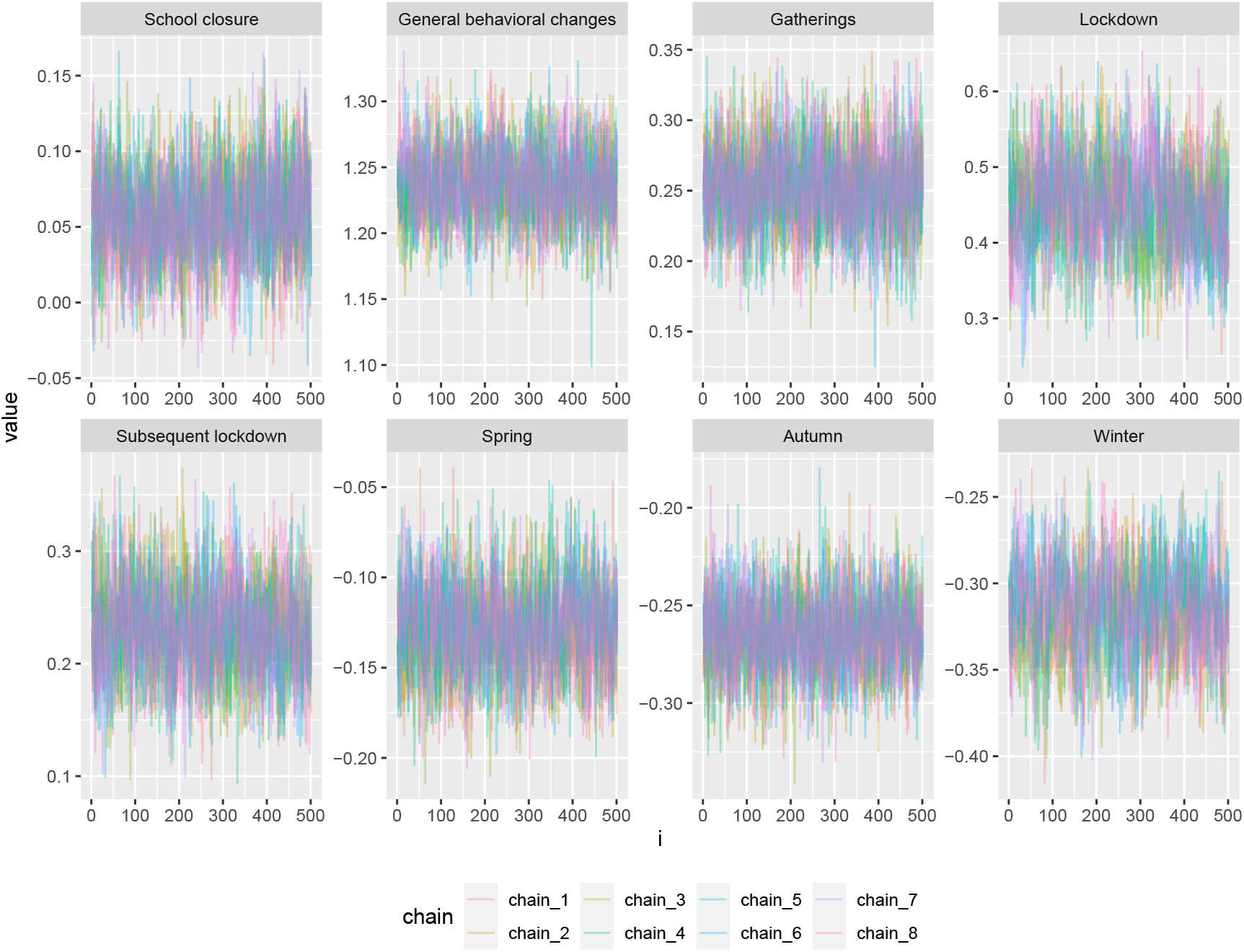
Traceplots of the mean effects of the NPIs. We use eight chains for each NPI and seasonal effect.

## Footnotes

1 Note that the decimal can be positive or negative depending on whether the calculated number in equation (2) is *>* 0.5 or not. We also want to mention that the quantity *C*_*t,m*_ can also contain infections which do not result in actual symptoms. Therefore *C*_*t,m*_ contains both, cases with actual symptoms and hypothetical cases without symptoms.

2 The data for the United Kingdom is not provided by the ECDC, however, the government provides data via an API: https://coronavirus.data.gov.uk/details/developers-guide

## References

Azmon, A., Faes, C., and Hens, N. (2014). On the estimation of the re-production number based on misreported epidemic data. Statistics in medicine, 33(7):1176–1192.

Baden, L. R., El Sahly, H. M., Essink, B., Kotloff, K., Frey, S., Novak, R., Diemert, D., Spector, S. A., Rouphael, N., Creech, C. B., et al. (2021). Efficacy and safety of the mrna-1273 sars-cov-2 vaccine. New England Journal of Medicine, 384(5):403–416.

Banholzer, N., van Weenen, E., Kratzwald, B., Seeliger, A., Tschernutter, D., Bottrighi, P., Cenedese, A., Salles, J. P., Vach, W., and Feuerriegel, S. (2020). Impact of non-pharmaceutical interventions on documented cases of covid-19. MedRxiv.

Bisoffi, Z., Pomari, E., Deiana, M., Piubelli, C., Ronzoni, N., Beltrame, A., Bertoli, G., Riccardi, N., Perandin, F., Formenti, F., et al. (2020). Sensitivity, specificity and predictive values of molecular and serological tests for covid-19: a longitudinal study in emergency room. Diagnostics, 10(9):669.

Brauner, J. M., Mindermann, S., Sharma, M., Johnston, D., Salvatier, J., Gavenčiak, T., Stephenson, A. B., Leech, G., Altman, G., Mikulik, V., et al. (2021). Inferring the effectiveness of government interventions against covid-19. Science, 371(6531).

Brooks, S., Gelman, A., Jones, G., and Meng, X.-L. (2011). Handbook of markov chain monte carlo. CRC press.

Brownstein, N. C. and Chen, Y. A. (2021). Predictive values, uncertainty, and interpretation of serology tests for the novel coronavirus. Scientific reports, 11(1):1–12.

Cohen, A. N. and Kessel, B. (2020). False positives in reverse transcription pcr testing for sars-cov-2. medRxiv.

Dehning, J., Zierenberg, J., Spitzner, F. P., Wibral, M., Neto, J. P., Wilczek, M., and Priesemann, V. (2020). Inferring change points in the spread of covid-19 reveals the effectiveness of interventions. Science.

Dong, E., Du, H., and Gardner, L. (2020). An interactive web-based dash-board to track covid-19 in real time. The Lancet infectious diseases, 20(5):533–534.

Fisman, D. N. and Tuite, A. R. (2021). Evaluation of the relative virulence of novel sars-cov-2 variants: a retrospective cohort study in ontario, canada. CMAJ: Canadian Medical Association Journal, 193(42):E1619.

Flaxman, S., Mishra, S., Gandy, A., Unwin, H., Coupland, H., Mellan, T., Zhu, H., Berah, T., Eaton, J., Perez Guzman, P., et al. (2020). Report 13: Estimating the number of infections and the impact of non-pharmaceutical interventions on covid-19 in 11 european countries.

Fraser, C., Donnelly, C. A., Cauchemez, S., Hanage, W. P., Van Kerkhove, M. D., Hollingsworth, T. D., Griffin, J., Baggaley, R. F., Jenkins, H. E., Lyons, E. J., et al. (2009). Pandemic potential of a strain of influenza a (h1n1): early findings. Science, 324(5934):1557–1561.

Gelman, A. and Rubin, D. B. (1992). Inference from iterative simulation using multiple sequences. Statistical science, 7(4):457–472.

Günther, F., Bender, A., Katz, K., Küchenhoff, H., and Höhle, M. (2021). Nowcasting the covid-19 pandemic in bavaria. Biometrical Journal, 63(3):490–502.

Haas, E. J., Angulo, F. J., McLaughlin, J. M., Anis, E., Singer, S. R., Khan, F., Brooks, N., Smaja, M., Mircus, G., Pan, K., et al. (2021). Impact and effectiveness of mrna bnt162b2 vaccine against sars-cov-2 infections and covid-19 cases, hospitalisations, and deaths following a nationwide vaccination campaign in israel: an observational study using national surveillance data. The Lancet, 397(10287):1819–1829.

Hale, T., Angrist, N., Goldszmidt, R., Kira, B., Petherick, A., Phillips, T., Webster, S., Cameron-Blake, E., Hallas, L., Majumdar, S., et al. (2021). A global panel database of pandemic policies (oxford covid-19 government response tracker). Nature Human Behaviour, 5(4):529–538.

Hall, V. J., Foulkes, S., Charlett, A., Atti, A., Monk, E. J., Simmons, R., Wellington, E., Cole, M. J., Saei, A., Oguti, B., et al. (2021). Sars-cov-2 infection rates of antibody-positive compared with antibody-negative health-care workers in england: a large, multicentre, prospective cohort study (siren). The Lancet, 397(10283):1459–1469.

Harris, C. R., Millman, K. J., van der Walt, S. J., Gommers, R., Virtanen, P., Cournapeau, D., Wieser, E., Taylor, J., Berg, S., Smith, N. J., Kern, R., Picus, M., Hoyer, S., van Kerkwijk, M. H., Brett, M., Haldane, A., Fernández del Río, J., Wiebe, M., Peterson, P., Gérard-Marchant, P., Sheppard, K., Reddy, T., Weckesser, W., Abbasi, H., Gohlke, C., and Oliphant, T. E. (2020). Array programming with NumPy. Nature, 585:357–362.

Hastings, W. K. (1970). Monte carlo sampling methods using markov chains and their applications. Biometrika, 57(1):97–109.

Islam, N., Sharp, S. J., Chowell, G., Shabnam, S., Kawachi, I., Lacey, B., Massaro, J. M., D’Agostino, R. B., and White, M. (2020). Physical distancing interventions and incidence of coronavirus disease 2019: natural experiment in 149 countries. bmj, 370.

Keehner, J., Horton, L. E., Pfeffer, M. A., Longhurst, C. A., Schooley, R. T., Currier, J. S., Abeles, S. R., and Torriani, F. J. (2021). Sars-cov-2 infection after vaccination in health care workers in california. New England Journal of Medicine, 384(18):1774–1775.

Khalili, M., Karamouzian, M., Nasiri, N., Javadi, S., Mirzazadeh, A., and Sharifi, H. (2020). Epidemiological characteristics of covid-19: a systematic review and meta-analysis. Epidemiology & Infection, 148.

Kumleben, N., Bhopal, R., Czypionka, T., Gruer, L., Kock, R., Stebbing, J., and Stigler, F. L. (2020). Test, test, test for covid-19 antibodies: the importance of sensitivity, specificity and predictive powers. Public Health, 185:88–90.

Lam, S. K., Pitrou, A., and Seibert, S. (2015). Numba: A llvm-based python jit compiler. In Proceedings of the Second Workshop on the LLVM Compiler Infrastructure in HPC, pages 1–6.

Li, Y., Campbell, H., Kulkarni, D., Harpur, A., Nundy, M., Wang, X., Nair, H., for COVID, U. N., et al. (2021). The temporal association of introducing and lifting non-pharmaceutical interventions with the time-varying reproduction number (r) of sars-cov-2: a modelling study across 131 countries. The Lancet Infectious Diseases, 21(2):193–202.

Mathieu, E., Ritchie, H., Ortiz-Ospina, E., Roser, M., Hasell, J., Appel, C., Giattino, C., and Rodés-Guirao, L. (2021). A global database of covid-19 vaccinations. Nature Human Behaviour, pages 1–7.

May, T. (2020). Lockdown-type measures look effective against covid-19. BMJ, 370.

O’Driscoll, M., Dos Santos, G. R., Wang, L., Cummings, D. A., Azman, A. S., Paireau, J., Fontanet, A., Cauchemez, S., and Salje, H. (2021). Age-specific mortality and immunity patterns of sars-cov-2. Nature, 590(7844):140–145.

Polack, F. P., Thomas, S. J., Kitchin, N., Absalon, J., Gurtman, A., Lockhart, S., Perez, J. L., Marc, G. P., Moreira, E. D., Zerbini, C., et al. (2020). Safety and efficacy of the bnt162b2 mrna covid-19 vaccine. New England Journal of Medicine.

Pritchard, E., Matthews, P. C., Stoesser, N., Eyre, D. W., Gethings, O., Vihta, K.-D., Jones, J., House, T., VanSteenHouse, H., Bell, I., et al. (2021). Impact of vaccination on new sars-cov-2 infections in the united kingdom. Nature Medicine, pages 1–9.

R Core Team (2021). R: A Language and Environment for Statistical Computing. R Foundation for Statistical Computing, Vienna, Austria.

Roberts, G. O. and Rosenthal, J. S. (2009). Examples of adaptive mcmc. Journal of Computational and Graphical Statistics, 18(2):349–367.

Santé Publique France (2021). Données relatives aux personnes vaccinées contre la covid-19. https://www.data.gouv.fr/fr/datasets/donnees-relatives-aux-personnes-vaccinees-contre-la-covid-19-1/ [Accessed: 2021-11-18].

Sharma, M., Mindermann, S., Rogers-Smith, C., Leech, G., Snodin, B., Ahuja, J., Sandbrink, J. B., Monrad, J. T., Altman, G., Dhaliwal, G., et al. (2021). Understanding the effectiveness of government interventions in europe’s second wave of covid-19. medRxiv.

Staerk, C., Wistuba, T., and Mayr, A. (2021). Estimating effective infection fatality rates during the course of the covid-19 pandemic in germany. BMC Public Health, 21(1):1–9.

Unwin, H. J. T., Mishra, S., Bradley, V. C., Gandy, A., Mellan, T. A., Coupland, H., Ish-Horowicz, J., Vollmer, M. A., Whittaker, C., Filippi, S. L., et al. (2020). State-level tracking of covid-19 in the united states. Nature communications, 11(1):1–9.

Virtanen, P., Gommers, R., Oliphant, T. E., Haberland, M., Reddy, T., Cournapeau, D., Burovski, E., Peterson, P., Weckesser, W., Bright, J., van der Walt, S. J., Brett, M., Wilson, J., Millman, K. J., Mayorov, N., Nelson, A. R. J., Jones, E., Kern, R., Larson, E., Carey, C. J., Polat, İ., Feng, Y., Moore, E. W., VanderPlas, J., Laxalde, D., Perktold, J., Cimrman, R., Henriksen, I., Quintero, E. A., Harris, C. R., Archibald, A. M., Ribeiro, A. H., Pedregosa, F., van Mulbregt, P., and SciPy 1.0 Contributors (2020). SciPy 1.0: Fundamental Algorithms for Scientific Computing in Python. Nature Methods, 17:261–272.

Voysey, M., Clemens, S. A. C., Madhi, S. A., Weckx, L. Y., Folegatti, P. M., Aley, P. K., Angus, B., Baillie, V. L., Barnabas, S. L., Bhorat, Q. E., et al. (2021). Safety and efficacy of the chadox1 ncov-19 vaccine (azd1222) against sars-cov-2: an interim analysis of four randomised controlled trials in brazil, south africa, and the uk. The Lancet, 397(10269):99–111.

Wallinga, J. and Lipsitch, M. (2007). How generation intervals shape the relationship between growth rates and reproductive numbers. Proceedings of the Royal Society B: Biological Sciences, 274(1609):599–604.

